# Intravenous methylphenidate for acute traumatic disorders of consciousness: A phase 1 dose-finding and target engagement study

**DOI:** 10.64898/2026.08.20.26359720

**Authors:** Brian L. Edlow, Megan E. Barra, David R. Schreier, Matteo Fecchio, Holly J. Freeman, Jian Li, Phoebe K. Lawrence, William R. Sanders, Anogue Meydan, Alexander S. Atalay, Maryam Masood, John E. Kirsch, Thomas P. Bleck, Joseph J. Fins, Joseph T. Giacino, Leigh R. Hochberg, Brian Healy, Ken Solt, Emery N. Brown, Yelena G. Bodien

## Abstract

**Background:** There are currently no therapies proven to accelerate recovery of consciousness for patients with acute severe traumatic brain injury (TBI) in the intensive care unit (ICU). Dopaminergic stimulation is a candidate strategy for reactivating subcortical networks that underly consciousness.

**Methods:** We performed an open-label, Phase 1 safety and dose-finding study of intravenous methylphenidate (IV MPH) in ICU patients with acute disorders of consciousness (DoC) caused by severe TBI. IV MPH, a potent and rapid-acting dopamine reuptake inhibitor, was administered in daily doses of 0.5, 1.0, and 2.0 mg/kg. The primary outcome measure was the number of adverse events (AEs) at each dose. IV MPH pharmacokinetics were measured for 24 hours after each dose. Pharmacodynamic target engagement – the effect of IV MPH on brain networks – was measured using EEG and resting-state functional MRI (rs-fMRI). A pharmacodynamic response was defined by change-point analysis of EEG and rs-fMRI time-series data. Behavioral responses were assessed using the Coma Recovery Scale-Revised (CRS-R).

**Results:** Between August 24, 2020, and April 1, 2024, we screened 488 ICU patients with TBI and enrolled 9 males (age 23-79 years) with acute traumatic DoC: coma (n=3), vegetative state/unresponsive wakefulness syndrome (n=3), and minimally conscious state (n=3). There were no serious AEs at any dose. Mild-moderate AEs observed at 1.0 mg/kg or 2.0 mg/kg included insomnia, emesis, paroxysmal sympathetic hyperactivity, and transaminitis. Maximum plasma MPH concentration ranged from mean (SD) 312.7 (100.6) ng/mL to 1319.5 (433.8) ng/mL and occurred within a median of 7-14 minutes across doses. Pharmacodynamic responses were observed via EEG in 7/8 participants who received 0.5 mg/kg (1/9 did not undergo EEG), 6/9 who received 1.0 mg/kg, and 4/6 who received 2.0 mg/kg. One of two patients who completed rs-fMRI showed a pharmacodynamic response. CRS-R level of arousal increased within 15 min of the IV MPH bolus for 6/9 participants at 0.5 mg/kg, 5/9 at 1.0 mg/kg, and 0/6 at 2.0 mg/kg.

**Conclusions:** For patients with acute severe TBI, IV MPH may be safe at daily doses of 0.5-2.0 mg/kg. EEG and rs-fMRI evidence of target engagement, accompanied by rapid behavioral increases in arousal, provides proof-of-principle that IV MPH reactivates subcortical networks underlying arousal, a prerequisite of consciousness. These findings support further evaluation of IV MPH in controlled trials.

## Introduction

For patients with acute severe traumatic brain injury (TBI), there are currently no therapies proven to promote recovery of consciousness in the intensive care unit (ICU).^1^ An intervention that accelerates recovery of consciousness in the ICU has the potential to improve the accuracy of prognostication^2^ and save lives by preventing premature withdrawal of life-sustaining treatment,^3^ the most common cause of in-hospital TBI-related deaths.^4^ Patients may also benefit by spending fewer days requiring mechanical ventilation and experiencing fewer ICU complications associated with immobilization, such as deep vein thrombosis and pneumonia.^5^ Moreover, a higher level of consciousness in the ICU may result in greater, and earlier, access to intensive inpatient rehabilitation.

Although a recent study provided proof-of-principle for the use of enteral and subcutaneous stimulants in ICU patients with acute brain injury,^6^ most previous efforts to promote recovery of consciousness in patients with severe TBI have largely focused on the subacute^7^ and chronic populations.^8,9^ Developing a treatment to promote consciousness for patients in the ICU is critical as decisions to withdraw life-sustaining treatment are often made within 72 hours, sometimes sooner.^4,10,11^ Of the many prognostic biomarkers that are used to guide these decisions, early recovery of consciousness has consistently been shown to be a primary determinant of long-term recovery^12–15^ and family decision-making.^16,17^ As such, a therapy that reveals a patient’s capacity for consciousness could substantially impact decisions about continuation of life-sustaining treatment in ICUs worldwide.^18^

To address this gap in clinical care, we performed a Phase 1 open-label, safety and dose-finding study of intravenous methylphenidate (IV MPH) in critically ill patients with acute disorders of consciousness (DoC) caused by severe TBI. MPH is a dopamine reuptake inhibitor that increases dopaminergic signaling from the ventral tegmental area in the midbrain to the diencephalon, basal forebrain, and cerebral cortex.^19–21^ MPH was selected for this trial based on evidence from animal models and proof-of-principle human studies that dopaminergic signaling may promote reemergence of consciousness after anesthesia^22–25^ and brain injury.^21,26,27^ The IV formulation of MPH was selected because of its faster onset (12-15 minutes in healthy conscious volunteers) and increased potency compared to the enteral formulation.^19^ In addition, the IV formulation may stimulate rapid phasic firing of neurons, which has the potential to promote recovery of cortical function, and hence consciousness, more so than the tonic neuronal firing observed with the enteral formulation of MPH.^19^

As this was a Phase 1 safety and dose-finding study, the primary outcome was the number of drug-related adverse events (AEs) at each dose of IV MPH. Secondary outcome measures pertained to pharmacokinetics (i.e., plasma half-life, time to peak plasma concentration, and maximum plasma concentration) and pharmacodynamics. For the pharmacodynamic analyses, we measured target engagement – the effect of IV MPH on brain activity – via electroencephalography (EEG) and resting-state functional MRI (rs-fMRI) biomarkers. We also performed serial standardized behavioral assessments using the Coma Recovery Scale-Revised (CRS-R).^28^ The study protocol included escalating daily doses of 0.5, 1.0, and 2.0 mg/kg IV MPH. We aimed to determine the safety profile and optimal dose of IV MPH for use in future clinical trials. By establishing the safety, pharmacokinetics, and target engagement of IV MPH, this first-in-human study provides the mechanistic and dosing foundation for future controlled efficacy trials.

## Materials and Methods

### Trial design and study participants

This Phase 1 trial – STIMPACT (<u>S</u>timulant <u>T</u>herapy Targeted to Individualized Connectivity <u>M</u>aps to <u>P</u>romote Re<u>ACT</u>ivation of Consciousness) – was approved by the United States Food and Drug Administration (FDA) in August, 2018 (IND 140675) and by the Mass General Brigham Institutional Review Board (IRB) on June 5, 2019. The study protocol was previously published,^29^ registered at ClinicalTrials.gov (NCT03814356) and approved by a Clinical Oversight Committee (COC) prior to study initiation on Aug 24, 2020. Throughout the trial, the COC met regularly to provide independent clinical guidance regarding participant eligibility, protocol implementation, and clinical safety issues. We also convened a Patient and Family Advisory Board, which reviewed the research protocol and expressed agreement with the aims and procedures. All methods, outcome measures, and statistical analyses have been previously described.^29^ Study launch and enrollment rate were impacted by supply chain disruptions, the COVID-19 pandemic, and mandatory implementation of updated workstreams to adhere to newly revised compounding requirements, as detailed in the Supplementary Methods.

We screened all patients with TBI admitted to the Neurosciences ICU at Massachusetts General Hospital (MGH) between August 24, 2020, and April 1, 2024. Screening and enrollment were expanded to the MGH Surgical ICU starting on May 3, 2022. Inclusion criteria were 1) age ≥ 18 years; and 2) DoC, defined as coma, vegetative state/unresponsive wakefulness syndrome (VS/UWS), or minimally conscious state (MCS), based on behavioral assessments performed by the clinical ICU team, and confirmed on the CRS-R after enrollment. Written, informed consent was provided by surrogate decision-makers. Enrollment could occur at any time during a patient’s ICU admission.

Study participation involved 5 consecutive days of assessments (**Figure 1**): baseline behavioral, EEG, and MRI assessments on Day 0, followed by four days of daily IV MPH at escalating doses: 0.5 mg/kg on Day 1, 1.0 mg/kg on Day 2, 2.0 mg/kg on Day 3, and the maximum tolerated dose on Day 4 during a rs-fMRI scan.

**Figure 1:**
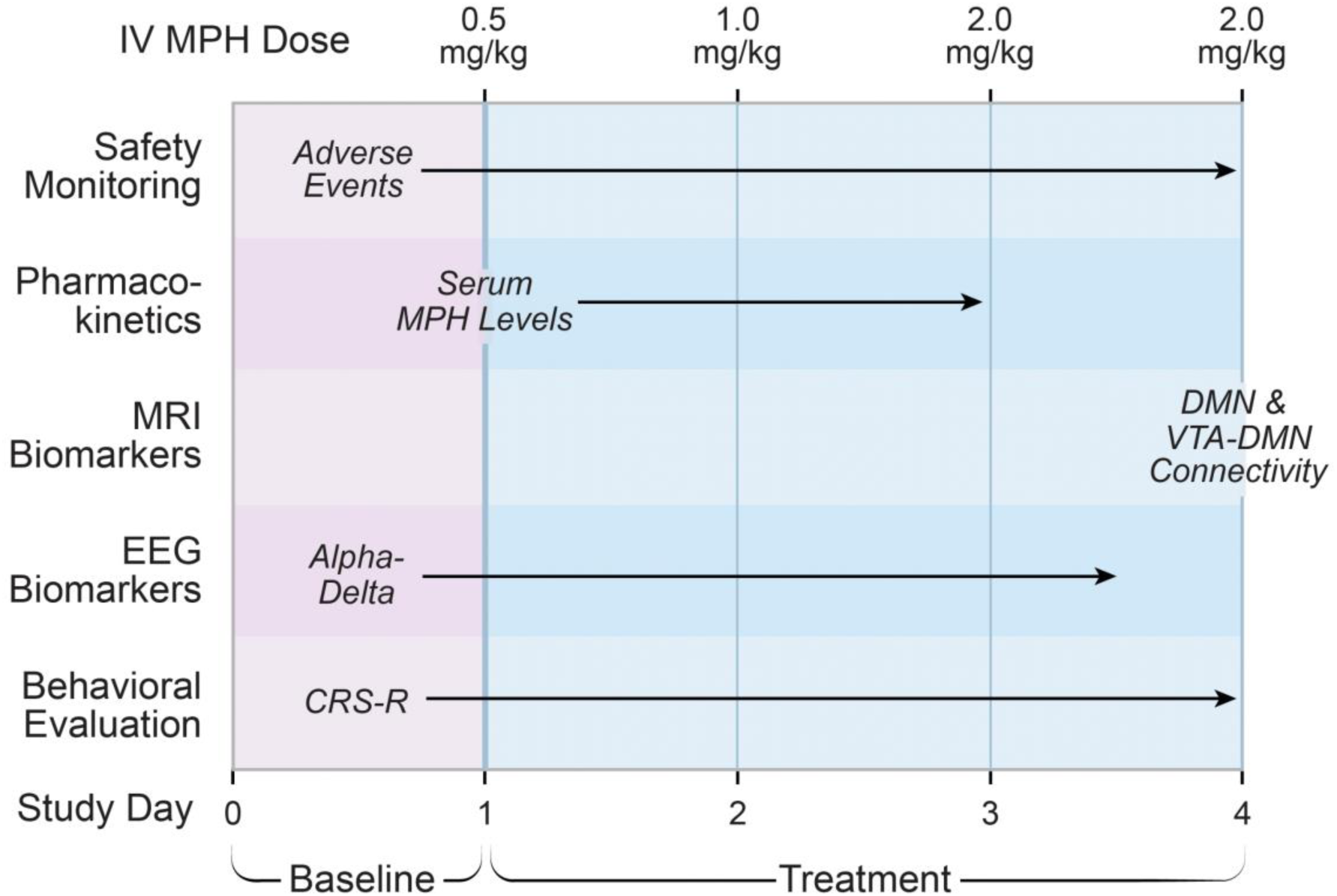
STIMPACT Study Protocol. Study participants received escalating daily doses of intravenous methylphenidate (IV MPH) for three days, followed by the highest tolerated dose on day 4. Abbreviations: CRS-R = Coma Recovery Scale-Revised; DMN = default mode network; EEG = electroencephalography; MRI = magnetic resonance imaging; VTA = ventral tegmental area.

We acquired EEG data continuously for four days: 24 hours prior to the first dose of IV MPH (Day 0), and three days during IV MPH dosing (EEG was discontinued on study protocol day 4 when participants underwent rs-fMRI). Participants who emerged from DoC during the 5-day study period, as defined by the behavioral criteria for emergence from MCS on the CRS-R assessment (i.e., functional object use or functional/accurate communication) remained in the study to undergo behavioral and EEG evaluations on the remaining study days, but did not receive additional doses of IV MPH.

### Procedures

IV MPH dosing was based on actual body weight. All doses were administered as a bolus at a rate of 20 mg/min. The primary outcome measure was the number of drug-related AEs at each of three IV MPH doses: 0.5, 1.0 and 2.0 mg/kg. Given the high rate of medical comorbidities for ICU patients with acute severe TBI,^5^ we used a checklist to track all clinical signs present before and after each dose.^7^ Only signs that were present after IV MPH dose administration, but not before, were considered AEs. The study team, in collaboration with ICU clinicians, tracked AEs for 24 hours after each dose (i.e., until the next dose, or until 24 hours after the final dose). If observed, AEs were then tracked until resolution. AEs and serious AEs (SAEs) were classified according to the Systematized Nomenclature of Medicine Clinical Terms (SNOMED CT) (https://www.snomed.org/). The study protocol stipulated that a participant would be withdrawn if an SAE was observed, or if an AE was observed at 0.5 mg/kg. If an AE was observed at 1.0 or 2.0 mg/kg doses, the participant continued in the study at the previously tolerated dose. All AEs were reported to surrogate-decision makers, the MGB IRB, the United States FDA, and the study COC. Any clinical sign that was newly observed after MPH dosing but did not meet prespecified criteria for an AE was recorded as an “event of clinical interest.” For example, the study COC determined at study initiation that agitation would be considered an event of clinical interest, not an AE, based on the frequency with which this clinical sign is observed in ICU patients recovering from acute severe TBI, independent of pharmacologic therapy.^5^

For pharmacokinetic analysis, serial plasma sampling was performed via arterial or central venous catheters at baseline (≤5 minutes before the bolus of IV MPH), and then at 5 mins (+/-5 mins), 15 mins (+/-5mins), 30 mins (+/-15 min), 1 hour (+/- 15 min), 1.5 hours (+/-15 min), 2 hours (+/- 15 min), 4 hours (+/-15 min), 8 hours (+/-15 min), 12 hours (+/-15 min) and 16 hours (+/-15 min) after bolus completion. Samples were stored at -80°C and shipped in batches for analysis to Worldwide Clinical Trials (Austin, TX, USA). Total MPH levels were quantified with liquid chromatographic/tandem mass spectrometry.

A standard 10-20 EEG montage with 19 electrodes, excluding the reference electrode located between Fz and Cz, was placed by a clinical EEG technician. Post-recording, we manually reviewed the EEG-synchronized video and audio recordings to verify the EEG timestamp for the initiation and completion of IV MPH bolus administration. We identified a continuous 5-min pre-bolus baseline, defined as the period after completion of the pre-bolus CRS-R assessment and before initiation of the bolus with the least amount of artifact and least interaction with the patient. Analyses included this time-period through the 15 minutes after completion of the IV MPH bolus. Additional details regarding EEG acquisition, pre-processing, and analysis are provided in the Supplementary Materials.

Rs-fMRI data were acquired using a blood-oxygen-level-dependent (BOLD) sequence with 2 mm isotropic resolution and a repetition time (TR) of 1.25 seconds. We recorded 10 minutes (482 frames) of rs-fMRI data pre-bolus and 30 minutes (1446 frames) post-bolus. Additional rs-fMRI sequence parameters have been previously described and are provided in the Supplementary Materials, along with details of rs-fMRI preprocessing.^15^ There was a brief (<1 minute) pause between the pre-bolus and post-bolus rs-fMRI acquisitions for logistical purposes. The start of the post-bolus rs-fMRI acquisition was precisely aligned with the start of the bolus itself, continuing through the ∼5-7 minute bolus and for ∼23-25 minutes after bolus completion. The timing of the rs-fMRI data acquisition was designed to capture IV MPH-evoked changes in brain dynamics occurring both during and after bolus administration.

Behavioral responses to IV MPH were assessed using the CRS-R on each day that IV MPH was administered: at 15 minutes before initiation of the dose as well as at 15-min and 60-min after the bolus was completed. The CRS-R arousal facilitation protocol was administered to promote wakefulness whenever sustained eye-closure was observed, and for participants with no eye-opening, prior to administration of each CRS-R item. On Day 4, the day of the rs-fMRI, the CRS-R was performed immediately prior to rs-fMRI, but it was not feasible to repeat the CRS-R while the patient was in the MRI scanner. Examiners recorded the level of consciousness in accordance with CRS-R protocol instructions: coma (no eye-opening during the examination and no evidence of conscious awareness), VS/UWS (eye-opening during the examination but no evidence of conscious awareness), MCS minus (MCS-; clear and reproducible signs of conscious awareness, such as visual pursuit, localization to noxious stimulation, but no evidence of language function), MCS plus (MCS+; clear and reproducible signs of conscious awareness with evidence of language function such as command-following or intelligible speech), or emergence from MCS (eMCS; evidence of functional object use or functional/accurate communication, but continued disorientation, significant cognitive impairment, agitation, or symptom fluctuation) as well as the CRS-R total score. Test Completion Codes were used to document the validity of the assessment (see Supplementary Materials). As this was an open-label study, the examiner was aware of the drug and dose being administered. CRS-R assessments were video recorded.

### Statistical Analysis

To determine the safety profile of each IV MPH dose, we estimated the proportion of participants who had AEs or SAEs and calculated the exact binomial 95% confidence interval. Our initial sample size calculation of n=22 was based on having a 90% chance of detecting any drug-related SAE that occurs with a frequency of ≥ 10% at any given dose.^29^ Our actual sample size of n=9 at 0.5 mg/kg and 1.0 mg/kg had a 90% chance of detecting any drug-related SAE that occurs with a frequency of ≥ 23%. Our sample size of n=6 at the 2.0 mg/kg dose provided a 90% chance of detecting any drug-related SAE that occurs with a frequency of ≥ 32%. Although one individual received 0.5 mg/kg dose twice and two received the 2.0 mg/kg dose twice, our primary analysis was conducted based on the number of participants receiving each dose once and not the number of times each dose was administered (i.e., n=9 for 0.5 mg/kg dose, even though 10 doses were administered). We also report AEs and SAEs by number of doses.

Pharmacokinetics were assessed for all subjects who received at least 1 dose and had at least 5 data points with quantifiable plasma concentration values. Pharmacokinetic parameters included maximum plasma concentration (C_max_), time to maximum concentration (T_max_), area under the concentration-time curve from time 0 to last quantifiable timepoint (AUC_0-T_), area under the concentration-time curve from time 0 to infinity (AUC_0-inf_), terminal half-life (t_1/2_) clearance (CL), and volume of distribution (Vd). Non-compartmental analysis was used to calculate pharmacokinetic parameters using Phoenix WinNonLin (Version 8.5.1; Certara, L.P; Princeton, NJ) with adjustment for the rate and duration of infusion (**Supplementary Figure 1**). Pharmacokinetic parameters were reported if the adjusted R^2^ value >0.80 or %AUC_extrap_ <20%. Concentrations below the lower limit of quantification were input as 0. Individual plasma concentrations of total MPH were listed and summarized by descriptive statistics.

To test for a pharmacodynamic response to IV MPH using EEG, we applied a changepoint analysis^30^ for each participant and study day that included an IV MPH bolus. Changepoint detection was based on the alpha-delta ratio (ADR) signal (mean ADR across channels) from the beginning of the pre-bolus baseline up to 15-min post-bolus completion. The window of observation for pharmacodynamic analyses is shown in **Supplementary Figure 1**. A participant was considered to be an MPH responder if the first EEG changepoint between initiation of the IV MPH bolus and 15 minutes post-bolus completion was positive (i.e., increase in the ADR following the changepoint relative to pre-bolus). A participant was considered to be an MPH non-responder if the first changepoint between initiation of the IV MPH bolus and 15 minutes post-bolus completion was negative or if there were no changepoints during this observation window. A negative changepoint represents a decrease in ADR following IV MPH. Given that this is the opposite effect expected in an IV MPH responder, participants with negative changepoints were classified as non-responders.

The rationale for limiting the changepoint analysis to 15 minutes post-bolus completion is that a behavioral assessment with the CRS-R was performed starting at post-bolus completion minute 15, and the CRS-R arousal facilitation protocol could confound an EEG analysis of the effects of IV MPH. We assessed for changepoints during IV MPH bolus administration based on prior studies suggesting that the rapid effects of IV MPH in the human brain could be observed within minutes of administration.^31^ To facilitate direct comparison of pre- and post-bolus EEG signals, we conducted a secondary analysis in which we compared the ADR between the 5-minute pre-bolus baseline and the period from the start of the IV MPH bolus to 15 min after IV MPH bolus completion. Methodologic details for this secondary analysis are provided in the Supplementary Materials.

We tested for a pharmacodynamic response to IV MPH using rs-fMRI using a similar approach to that of EEG. In the primary analysis, we concatenated the pre- and post-bolus data into one time series and computed dynamic functional connectivity between the VTA and cortical default mode network (DMN) nodes (VTA-DMN connectivity) using a sliding-window-based Pearson correlation as prespecified in the study protocol.^29^ We varied the duration of the sliding window from 30s to 60s and averaged the connectivity results to minimize any impacts from the arbitrary choice of the sliding window size.^32^ We then performed a changepoint analysis on the resulting VTA-DMN connectivity time series using the same changepoint detection algorithm used for the EEG dataset. A changepoint was detected if the mean squared error was reduced by at least 10% of the total variance of the connectivity time series. The results were not sensitive to the choice of this threshold (5%-30%). Analyses were conducted using customized Matlab code.

MPH responders were defined using the same criteria as for EEG (i.e., the first changepoint after initiation of the IV MPH bolus being positive). In the secondary analysis, we performed a static functional connectivity analysis using the seed-based connectivity (SBC) pipeline from the CONN Functional Connectivity Toolbox^33^, as detailed in the Supplementary Materials. We compared VTA-DMN connectivity and intra-network DMN connectivity (i.e., connectivity between DMN cortical nodes) during the 10-minute pre-bolus time interval (baseline) and during two 10-minute post-bolus time intervals: from minutes 5 to 15, and from minutes 10 to 20, allowing for up to 1 minute of adjustment of the time windows if motion artifacts were observed in the rs-fMRI data (e.g., in P8).

## Results

We screened 488 patients admitted to the Neurosciences and Surgical ICUs at MGH with a TBI, of whom 474 were excluded. Surrogates of 4 eligible participants declined participation. We enrolled 10 patients during the study period. One surrogate decision-maker withdrew consent prior to initiation of study procedures, yielding a final cohort of 9 participants (age range 23 to 79 years; all male). A Consort Diagram is provided in **Supplementary Figure 2**. The mechanisms of TBI were motor vehicle accident (n=4), fall (n=3) and assault (n=2). Additional details regarding patient demographics and clinical characteristics are provided in **Table 1**. Details regarding baseline behavioral, MRI, and EEG assessments on study Day 0 are provided in **Supplementary Table 1**. Continuous EEG was performed in all 9 patients and rs-fMRI in 2. The first dose of IV MPH was administered on median day 10 (range = 5 to 33 days) post-injury. Weight-based doses ranged from 27.5 to 61.3 mg at 0.5mg/kg, 55.0 to 123.0 mg at 1.0 mg/kg, and 139.0 to 246.0 mg at 2.0 mg/kg. Dosing data for individual subjects is summarized in **Supplementary Table 2**.

**Table 1:** Patient Demographics, Clinical Characteristics, and Timing of Intravenous Methylphenidate. The initial GCS (iGCS) is defined as the best (i.e. highest) and worst (i.e. lowest) post-resuscitation GCS score assessed by a qualified clinician who performed a reliable examination (not confounded by sedation and/or paralytics) prior to ICU admission. LoC is assessed via behavioral evaluation with the CRS-R as coma, VS, MCS-, MCS+, or eMCS (emerged from MCS but disoriented). For the LoC Before the First Dose of IV MPH, we report the CRS-R-derived LoC immediately before the first dose of IV MPH was administered. For the LoC After the Last Dose of IV MPH, we report the highest LoC at the 15-minute or 60-minute CRS-R performed after the last dose of IV MPH that was administered in the ICU (post-dose CRS-R assessments could not be performed while patients were in the MRI scanner). Abbreviations: CKD = Chronic Kidney Disease, CRS-R= Coma Recovery Scale-Revised; eMCS = emerged from minimally conscious state; F = female; GCS = Glasgow Coma Scale; HL = hyperlipidemia HTN = hypertension, IV MPH = intravenous methylphenidate; LoC = Level of Consciousness immediately prior to first dose of IV MPH; M = male; MCS-= minimally conscious state without language; MCS+ = minimally conscious state with language; MVA = motor vehicle accident; N/A = not applicable; Ped = pedestrian; TBI = traumatic brain injury; VS = vegetative state. * Patient seizing at time Emergency Medical Services arrived at crash scene. Required high doses of benzodiazepines to stop seizures. Thus, no examination performed in the absence of seizure or sedatives prior to admission to intensive care unit.

| ID | Age Range (yrs) | Sex | Medical History | TBI Mechanism | iGCS | Post-injury Day of First IV MPH Dose | Days of IV MPH Therapy | IV MPH Doses Received (mg/kg) | LoC Before First Dose of IV MPH | LoC After Last Dose of IV MPH |
| --- | --- | --- | --- | --- | --- | --- | --- | --- | --- | --- |
| P1 | 40-49 | M | Polysubstance abuse | Assault | 3-9T | 6 | 2 | 0.5, 1.0 | MCS+ | eMCS |
| P2 | 20-29 | M | None | MVA | 3-6 | 9 | 3 | 0.5, 1.0, 2.0 | MCS- | MCS+ |
| P3 | 40-49 | M | Hepatocellular carcinoma, meningioma | MVA | 3-4 | 6 | 3 | 0.5, 1.0, 2.0 | VS/UWS | MCS+ |
| P4 | 70-79 | M | HTN, HL, ischemic stroke (R basal ganglia), Stage 3 CKD, depression, Bell's palsy | Fall | 4T-15 | 5 | 4 | 0.5, 1.0, 2.0, 2.0 | Coma | VS/UWS |
| P5 | 50-59 | M | HTN, Polysubstance abuse | Assault | 3-6 | 10 | 2 | 0.5, 1.0 | MCS- | MCS- |
| P6 | 70-79 | M | HTN, atrial fibrillation | Fall down 10 stairs | 10-12 | 10 | 3 | 0.5, 1.0, 2.0 | VS/UWS | MCS- |
| P7 | 20-29 | M | None | MVA | 3-4T | 33 | 3 | 0.5, 1.0, 2.0 | VS/UWS | Coma |
| P8 | 60-69 | M | None | Fall down 12 stairs | 3-3T | 13 | 4 | 0.5, 1.0, 2.0, 2.0 | Coma | VS/UWS |
| P9 | 20-29 | M | None | MVA | 3-3T* | 22 | 3 | 0.5,1.0, 0.5 | Coma | VS/UWS |

**Table 2:** Adverse Events. Abbreviations: AE = adverse event; ALT = alanine transaminase; AST = aspartate aminotransferase; IV = intravenous; N/A = not applicable; PEG = percutaneous endoscopic gastrostomy.

| ID | AE Type | Post-TBI Day # | MPH to AE Onset (hours) | AE Duration | Severity | Expectedness | Relatedness | Therapies Administered | Implications for Subsequent MPH Dosing |
| --- | --- | --- | --- | --- | --- | --- | --- | --- | --- |
| 0.5 mg/kg |  |  |  |  |  |  |  |  |  |
| N/A |  |  |  |  |  |  |  |  |  |
| 1.0 mg/kg |  |  |  |  |  |  |  |  |  |
| P1 | Insomnia | 7 | 11 | 5 hours | Moderate | Expected | Related | Melatonin 5 mg<br>PNG<br>Lorazepam 2mg<br>IV x2 | Not applicable. Patient had emerged from MCS to eMCS and therefore no longer eligible to receive IV MPH. |
| P9 | Paroxysmal sympathetic hyperactivity | 23 | 0 | 1 hour | Moderate | Expected | Related | Dexmedetomidine 0.3<br>Propofol 20 | Yes – down to 0.5 mg/kg on next day |
| P9 | Emesis | 23 | 1 | Seconds | Mild | Expected | Related | None | Yes – down to 0.5 mg/kg on next day |
| 2.0 mg/kg |  |  |  |  |  |  |  |  |  |
| P4 | Increased ALT/AST | 8 | ~24 | Persisted for 6 days, until goals of care transitioned to comfort-focused care, at which time labs were no longer checked. | Moderate (grade 2) | Expected | Related | None | None, because AE occurred on the day after study completion (i.e., the ALT did not increase to 3x the upper limit of normal until post-TBI Day 9) |

A cumulative total of four AEs was recorded in 3 patients. All AEs were expected and related to the IV MPH, per FDA definitions and based on consensus opinion of the CoC. At the 0.5 mg/kg dose, we observed 0 AEs (10 doses in 9 patients). At 1.0 mg/kg there were 3 AEs out of 9 doses, which were observed in 2 patients (proportion 2/9; 95% CI [0.03 – 0.60]). At 2.0 mg/kg there was 1 AE out of 8 doses, given to 6 patients (proportion 1/6; 95% CI [0.004, 0.64]). There were no SAEs at 0.5 mg/kg (estimated proportion 0/9; 95% CI [0, 0.34]), 1.0 mg/kg (estimated proportion 0/9; 95% CI [0, 0.34]), or 2.0 mg/kg (estimated proportion 0/6; 95% CI [0, 0.46]). When considering the administration of multiple doses to the same participant, there were 0 SAEs at 0.5 mg/kg (estimated proportion 0/10; 95% CI [0, 0.31]) and 2.0 mg/kg (estimated proportion 0/8; 95% CI [0, 0.37]).

Among the 3 participants for whom 4 AEs were observed, one (P9) received the previously tolerated dose (2 AEs at 1.0 mg/kg then received 0.5mg/kg the next day); one (P1) did not receive another dose because of emergence from MCS; and one (P4) experienced an AE after receiving all doses of IV MPH, including two 2.0 mg/kg doses, and hence did not miss any doses of the medication. Details summarizing the reasons for missed doses are provided in **Supplementary Table 3.**

Events of clinical interest that did not meet criteria for an AE are summarized in **Supplementary Table 4**. These included agitation/restlessness (n=2 participants), diaphoresis (n=1 participant, who experienced diaphoresis at all three doses); and increased alanine transaminase (ALT)/aspartate aminotransferase (AST) levels (n=1 participant). With respect to the latter, participant P4 experienced elevated ALT/AST that did not meet criteria for an AE on study days 1, 2 and 3, but met criteria for an AE on day 4.

At the 0.5 mg/kg dose, 4/9 participants received pre-MPH treatment with intravenous cardiovascular medications to lower their heart rate or blood pressure, and 7/9 participants received post-MPH cardiovascular medications to maintain their heart rate and blood pressure within prespecified targets. Utilization rates of pre-MPH and post-MPH cardiovascular medications were lower at the 1.0 and 2.0 mg/kg doses. Details regarding pre-MPH and post-MPH cardiovascular medications, including which medications were administered for clinical indications versus for compliance with the study protocol, are provided in **Supplementary Table 5**.

Pharmacokinetic analyses were completed for 8 participants who received 0.5 mg/kg, 8 who received 1.0 mg/kg, and 5 who received 2.0 mg/kg IV MPH. P2 was excluded from pharmacokinetic analysis because blood samples were obtained from a central venous catheter through which the MPH bolus was infused, producing inaccurate results. The median T_max_ for the 0.5 mg/kg, 1.0 mg/kg, and 2.0 mg/kg doses was 0.12 hours, 0.15 hours, and 0.23 hours, respectively. A dose-proportional increase in total MPH plasma concentrations was observed for C_max_ with a mean (SD) of 312.72 (100.62) ng/mL, 676.91 (309.16) ng/mL, and 1319.53 (433.76) ng/mL following administration of 0.5, 1.0, and 2.0 mg/kg IV MPH, respectively. Mean T_½_ was similar across dose cohorts: 5.07, 4.39, and 5.01 hours following the 0.5 mg/kg, 1.0 mg/kg, and 2.0 mg/kg doses, respectively. Inter-subject variability in terminal elimination half-life was observed. Across patients, T_½_ varied from a minimum of 2.44 hours to a maximum of 9.34 hours. A summary of plasma concentration curves and pharmacokinetic measures is provided in **Figure 2** and **Table 3**. Individual PK parameters stratified by dose are provided in **Supplementary Table 6**.

**Figure 2:**
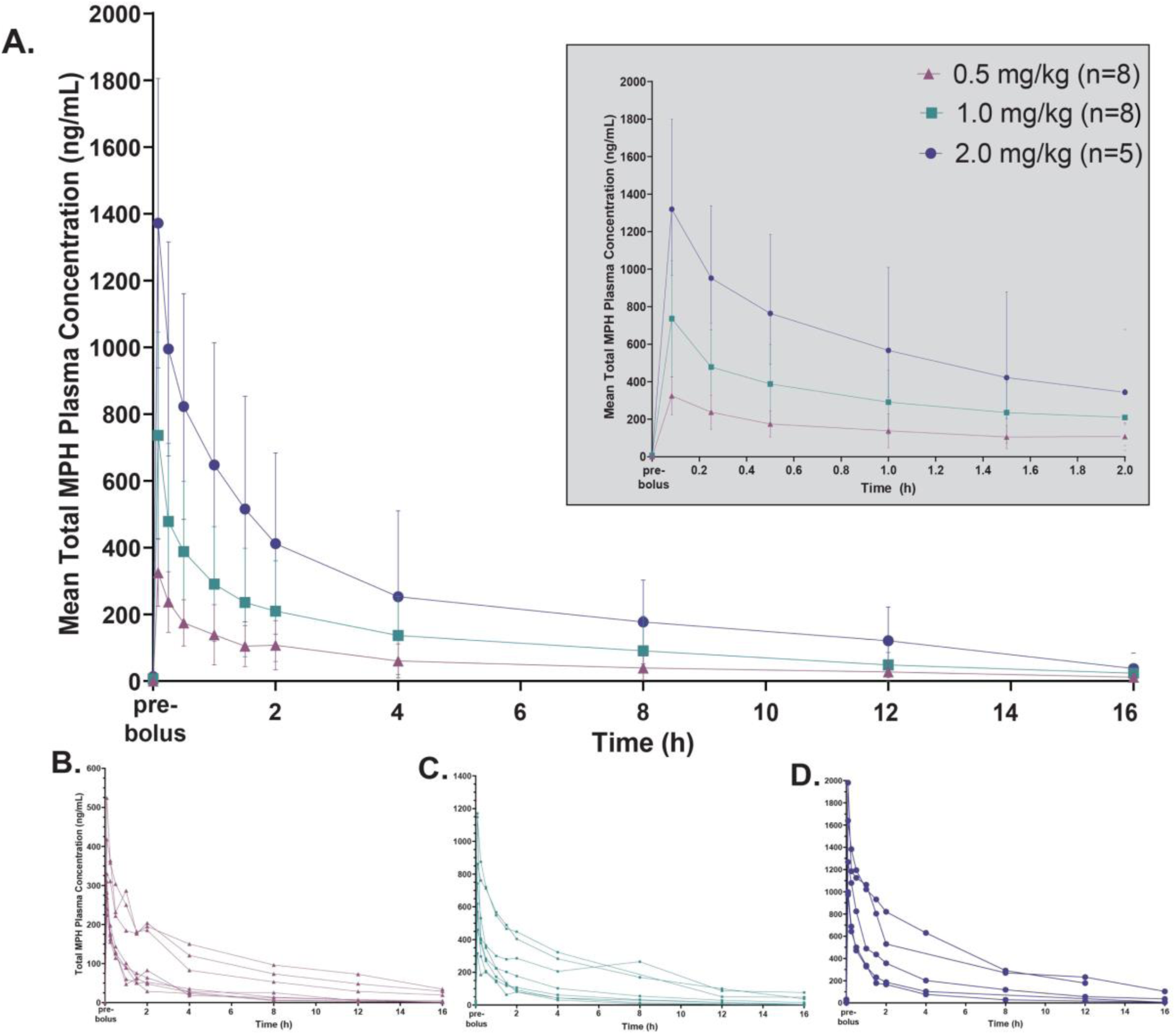
Total Methylphenidate Plasma Concentrations Following Intravenous Administration. Total methylphenidate (MPH) concentration (ng/ml) was measured via blood samples drawn at baseline (≤ 5 minutes before the bolus of IV MPH), and then at 5 mins (+/- 5 mins), 15 mins (+/- 5mins), 30 mins (+/- 15 min), 1 hour (+/- 15 min), 1.5 hours (+/- 15 min), 2 hours (+/- 15 min), 4 hours (+/- 15 min), 8 hours (+/- 15 min), 12 hours (+/- 15 min) and 16 hours (+/- 15 min) after bolus completion. Mean MPH plasma concentrations are shown in (A) for the 0.5 mg/kg, 1.0 mg/kg and 2.0 mg/kg doses, with the grey inset showing concentration curves for the first two hours after each bolus. Patient-specific concentration curves are shown for the 0.5 mg/kg dose in (B), for the 1.0 mg/kg dose in (C), and for the 2.0 mg/kg dose in (D).

**Table 3.**
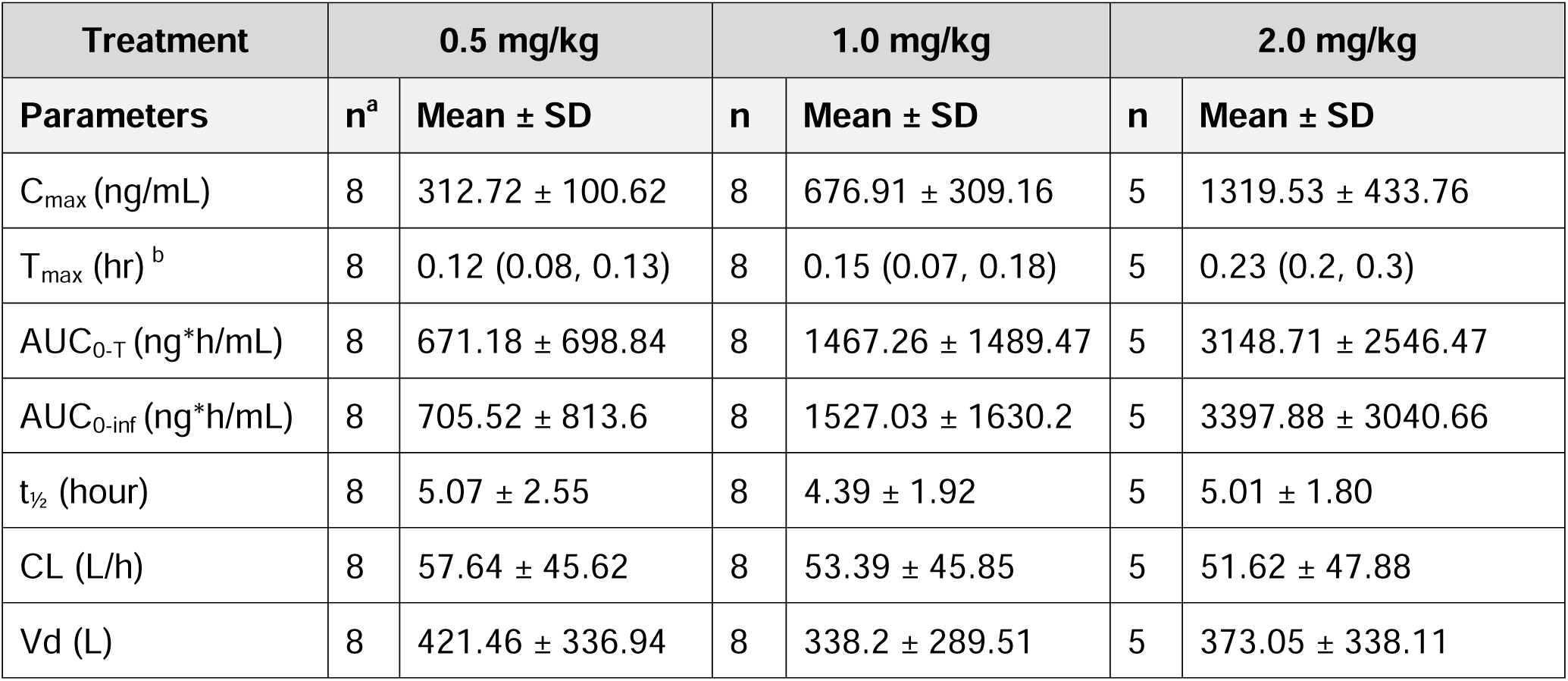
Plasma Pharmacokinetic Parameters of Total Methylphenidate after Intravenous Doses of Methylphenidate. All data are presented as the geometric mean and standard deviation. Abbreviations: AUC_0-inf_ = area under the concentration-time curve from time 0 after bolus initiation to infinity; AUC_0-t_= area under the concentration-time curve from time 0 after bolus initiation to last quantifiable timepoint; CL = total body clearance; Cmax = maximum plasma concentration; SD = standard deviation; t_1/2_= terminal elimination half-life; T_max_= time from bolus initiation to maximum concentration; Vd = volume of distribution. ^a^One subject (STIM_002) was excluded from the pharmacokinetic and statistical analysis for inaccurate sampling collection. ^b^ T_max_ reported as median (minimum, maximum).

In the primary EEG pharmacodynamic analyses, we observed an ADR signal-related positive changepoint at the 0.5 mg/kg, 1.0 mg/kg, and 2.0 mg/kg dose for 87.5%, 66.7%, and 66.7% of participants respectively (**Figure 3**). Therefore, at each dose, most participants were classified as IV MPH responders. Of note, the single non-responder at 0.5 mg/kg (P6) demonstrated changes in alpha and delta power that began during the bolus and were sustained until the 15-minute CRS-R assessment. However, these changes, which are visible on the EEG spectrogram, did not meet the prespecified criteria for an IV MPH responder. A summary of EEG changepoints for each participant on each day of IV MPH administration is provided in the Supplement (**Supplementary Figure 3**). EEG data for one participant (P5) was not available during IV MPH administration at the 0.5 mg/kg dose because of technical difficulties with EEG lead placement.

**Figure 3:**
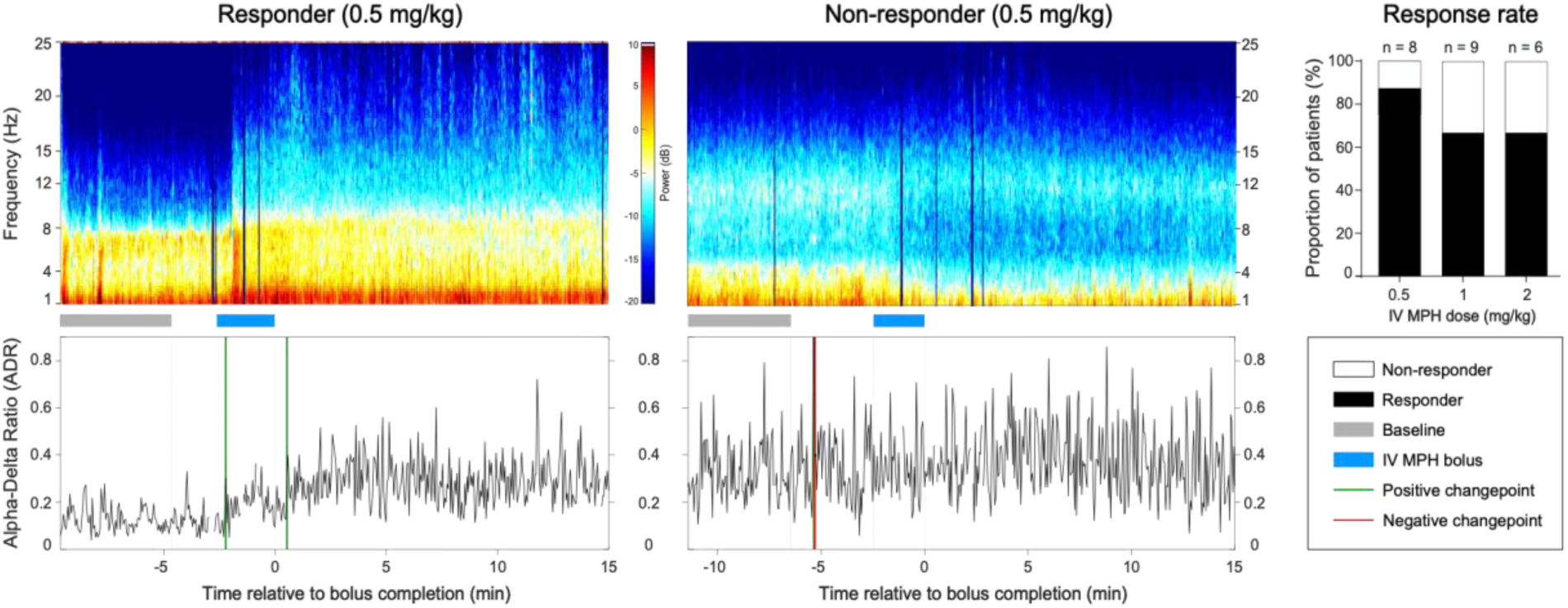
Electroencephalography Pharmacodynamic Responses to Intravenous Methylphenidate. Electroencephalography (EEG) spectrograms (top row) and alpha-delta ratio plots (bottom row) are shown for a representative responder (P2) and non-responder (P6). The dark blue vertical bars in the spectrograms indicate time periods for which EEG data were excluded because of artifacts. The proportion of patients who experienced a pharmacodynamic response on EEG at each dose – 0.5 mg/kg, 1.0 mg/kg and 2.0 mg/kg – is shown in the top right panel.

In the secondary EEG pharmacodynamic analysis, the ADR significantly increased in 5/8 participants (62.5%) after bolus initiation, compared to the baseline period at 0.5 mg/kg (**Table 4**). The proportion of participants with a post-bolus increase in ADR was 55.6% (5/9) and 33.3% (2/6) at the 1.0 and 2.0 mg/kg doses, respectively. There was agreement between the changepoint-based (primary analysis) and the bootstrap-based (secondary analysis) results for 4/8 at 0.5 mg/kg, 6/9 at 1.0 mg/kg, and 4/6 at 2.0 mg/kg (Table 4). Sedative medications administered within 1 hour of the IV MPH bolus are summarized in **Supplementary Table 7**.

**Table 4.**
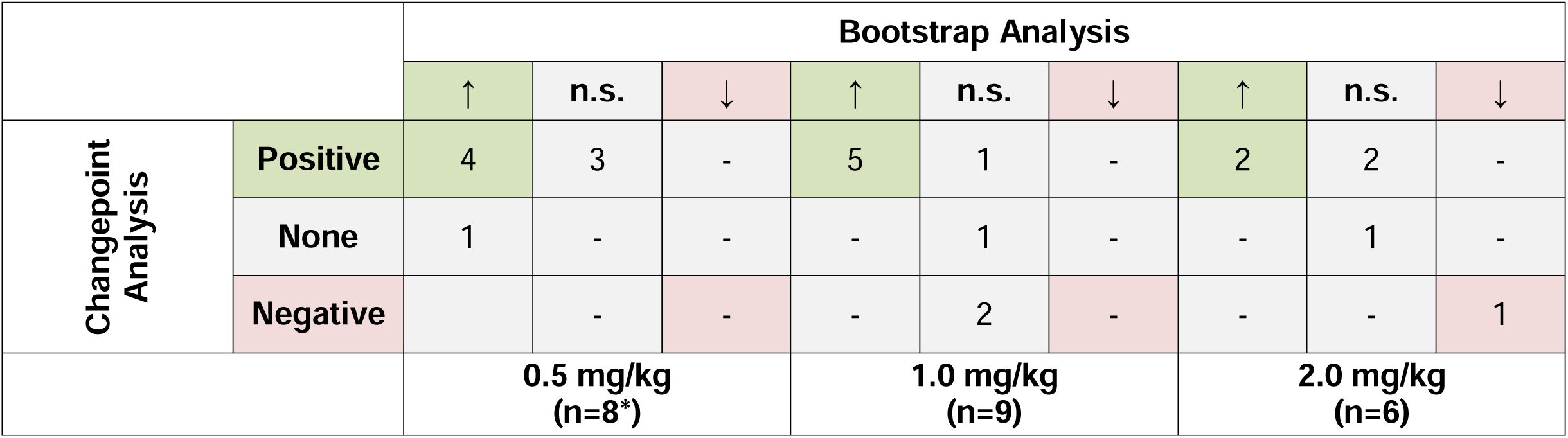
Association between Changepoint and Bootstrap Alpha-Delta Ratio Analyses. Changepoint (CP) responses detected via electroencephalography (EEG) were assessed during the period from the start of the intravenous methylphenidate (IV MPH) bolus to 15 min following bolus completion. ↑ indicates a significant increase and ↓ a significant decrease of the alpha-delta ratio (ADR) from the 5-min baseline to the period from the start of the IV MPH bolus to 15 min following bolus completion. Abbreviations: n.s. = no significant change. *EEG data for one participant (P5) was not available during IV MPH administration at the 0.5 mg/kg dose because of technical difficulties with EEG lead placement.

|  |  | Bootstrap Analysis |  |  |  |  |  |  |  |  |
| --- | --- | --- | --- | --- | --- | --- | --- | --- | --- | --- |
|  |  | ↑ | n.s. | ↓ | ↑ | n.s. | ↓ | ↑ | n.s. | ↓ |
| Changepoint Analysis | Positive | 4 | 3 | - | 5 | 1 | - | 2 | 2 | - |
|  | None | 1 | - | - | - | 1 | - | - | 1 | - |
|  | Negative |  | - | - | - | 2 | - | - | - | 1 |
|  |  | 0.5 mg/kg<br>(n=8*) |  |  | 1.0 mg/kg<br>(n=9) |  |  | 2.0 mg/kg<br>(n=6) |  |  |

In the rs-fMRI pharmacodynamic analysis, we identified one IV MPH responder (P4) and one non-responder (P8). The responder (P4) had one positive changepoint within a minute of bolus completion (∼7 mins after bolus initiation). This increase in connectivity was sustained throughout the remaining recording period. The non-responder (P8) had no change points and the connectivity fluctuated around a Pearson’s correlation of zero (**Figure 4A**). The remaining participants did not complete the rs-fMRI component of the study due to emergence from MCS (n=1), agitation (n=1), medical instability or comorbidities (n=3), body metal (n=1), and change in goals of care to comfort-focused care (n=1), as detailed in Supplementary Table 3.

**Figure 4:**
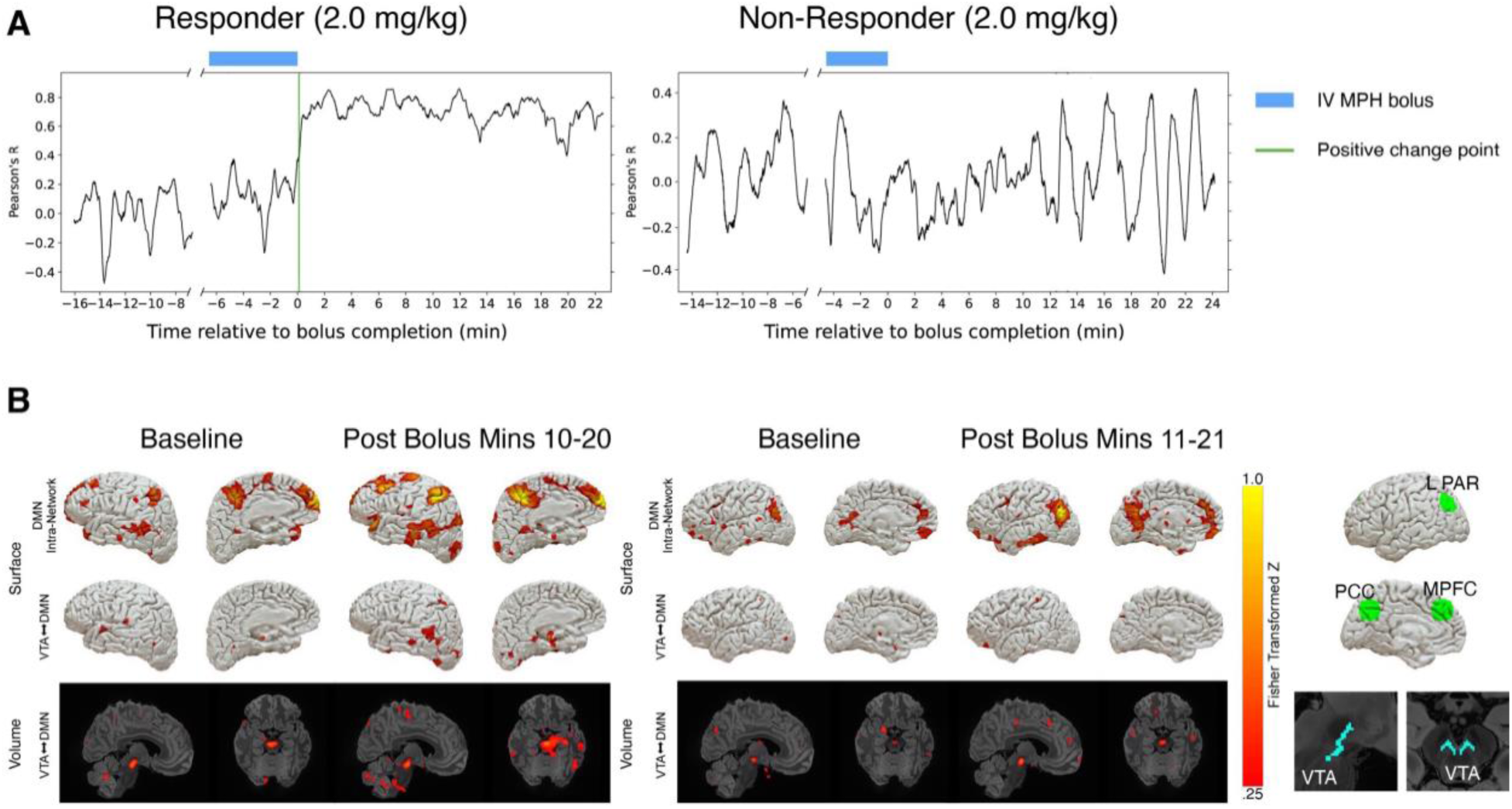
Resting-state Functional MRI Pharmacodynamic Responses to Intravenous Methylphenidate. (A) Dynamic resting-state functional connectivity data are shown for a responder (P4) and non-responder (P8) with bolus administration period and change points marked along the time axis. (B) Comparison of the seed-based functional connectivity (z map) between the baseline and post-bolus rs-fMRI is shown for the same two patients. The first row shows intra-network default mode network (DMN) connectivity plotted on the cortical surface. The second and third row show ventral tegmental area (VTA) connectivity with the DMN (VTA-DMN) plotted on the surface and volume, respectively. The right-most panel illustrate the VTA and DMN ROIs used for this analysis.

In the seed-based ROI analysis (secondary analysis), P4 experienced increases in functional connectivity following the bolus relative to baseline (**Figure 4B**; all units as Fisher-transformed Z scores). Intra-network DMN connectivity increased from 0.48 at baseline to 0.53 (post-bolus minutes 5–15) and remained elevated at 0.55 (post-bolus minutes 10–20). VTA–DMN connectivity increased from 0.01 to 0.14 and remained elevated at 0.16, respectively, across the same time windows. For P8, DMN connectivity increased from 0.32 at baseline to 0.50 (post-bolus minutes 6–16) and 0.55 (post-bolus minutes 11–21). P4 was also classified as a responder on EEG at all doses of IV MPH. P8 was a responder on EEG at the 0.5 mg/kg dose, but not at the 1.0 or 2.0 mg/kg doses.

Behavioral data are summarized in **Figure 5**. All behavioral data for the CRS-R assessments performed 15 minutes pre-dose, 15 minutes post-dose and 60 minutes post-dose are provided in **Supplementary Tables 8, 9, and 10**. The level of consciousness immediately prior to the first dose of IV MPH (based on the CRS-R examination) was coma (n=3), VS/UWS (n=3), MCS- (n=2) and MCS+ (n=1). The most noticeable post-bolus behavioral observation was a rapid improvement in level of arousal (as demonstrated by an increase in the CRS-R Arousal Subscale score 15- minutes post-bolus) for 6/9 (67%) participants at 0.5 mg/kg and 5/8 (63%) at 1.0 mg/kg. CRS-R arousal scores did not improve for any patient (0/6; 0%) at 2.0 mg/kg. An improvement in CRS-R total scores within 15 minutes was observed for 7/9 (77%) participants at 0.5 mg/kg, 5/9 (56%) participants at 1.0 mg/kg, and 3/6 (50%) participants at 2.0 mg/kg. CRS-R-derived DoC diagnostic categories improved within 15 minutes for 3/9 (33%) participants at 0.5 mg/kg, 1/9 (11%) at 1.0 mg/kg, and 0/6 (0%) at 2.0 mg/kg. Behavioral changes had variable duration, with some no longer present by 60 minutes post-bolus. Of note, the high rate of pharmacodynamic responses observed in the primary EEG analysis (87.5%, 66.7%, and 66.7% at the 0.5, 1.0 and 2.0 mg/kg doses, respectively), generally exceeded the rate of improvement in CRS-R arousal scores, CRS-R-derived diagnostic category, and CRS-R total scores. A summary of pharmacodynamic and behavioral responses for each individual participant is provided in **Supplementary Table 11**.

**Figure 5:**
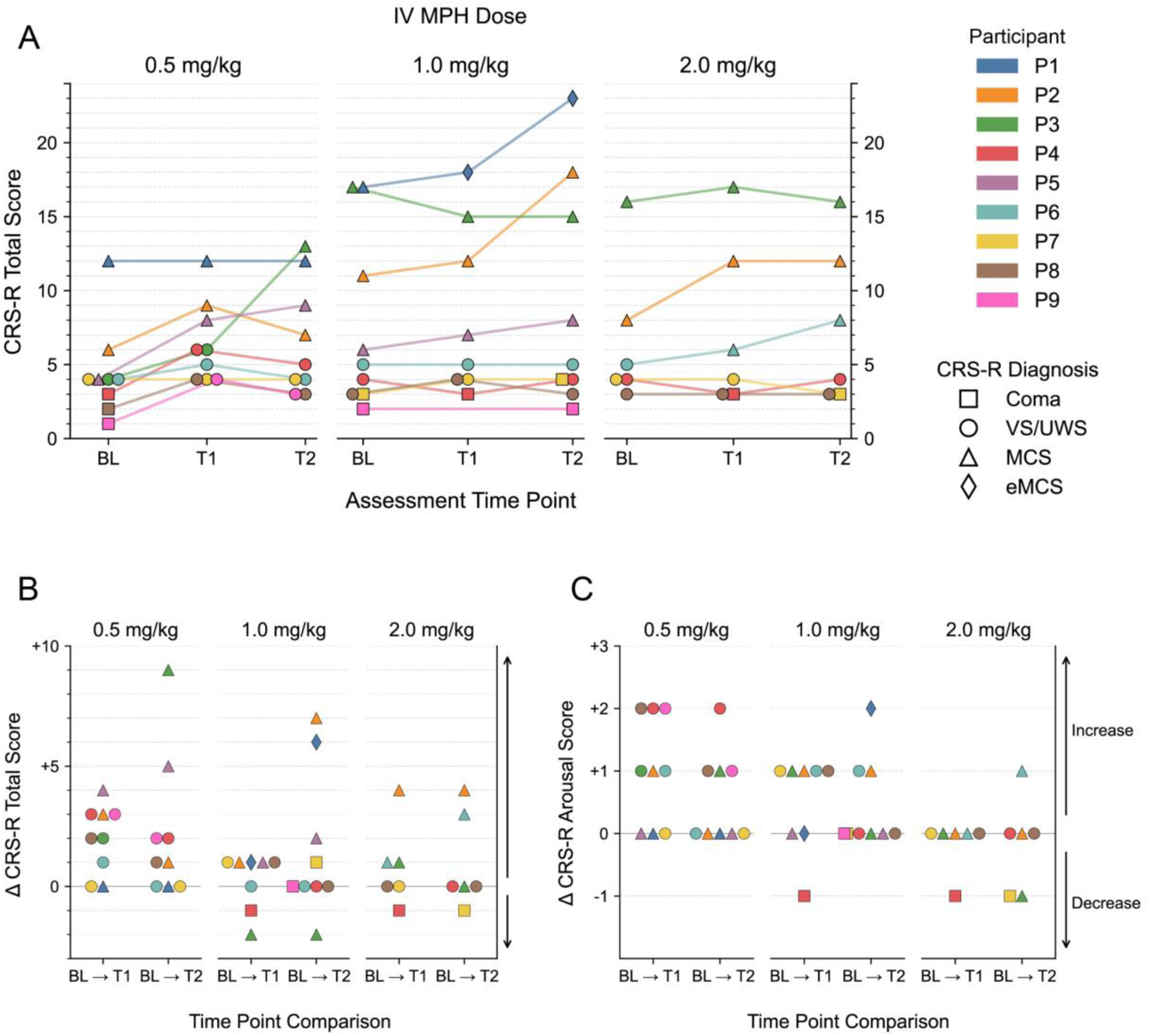
Behavioral Observations Following Intravenous Methylphenidate. Behavioral changes in level of consciousness were assessed via the Coma Recovery Scale-Revised (CRS-R), as shown in the top row. Longitudinal changes in level of consciousness are shown as changes in CRS-R Total Score (middle row) and longitudinal changes in level of arousal are shown as changes in the CRS-R Arousal Subscale Score (bottom row). Abbreviations: BL = baseline; eMCS = emergence from minimally conscious sate; MCS = minimally conscious state; T1 = timepoint 1 (i.e., 15 minutes post-bolus); T2 = timepoint 2 (i.e., 60 minutes post-bolus); VS/UWS = vegetative state/unresponsive wakefulness syndrome.

## Discussion

Our results provide preliminary evidence that IV MPH is safe to administer at doses of 0.5 to 2.0 mg/kg to patients with acute DoC following severe TBI. There were no SAEs, though mild-moderate drug-related AEs were observed at the 1.0 and 2.0 mg/kg doses, indicating that 0.5 mg/kg may be the safest dose of IV MPH in this population. IV MPH reached peak concentration in blood plasma at a median of 12-15 minutes (range 4 – 18 minutes, across doses) after administration and, in some participants, appeared to rapidly affect brain function, as assessed by EEG, rs-fMRI, and standardized behavioral assessments. Acknowledging that behavioral assessments may have been confounded by the open-label study design, IV MPH was frequently associated with rapid increases in behavioral signs of arousal (i.e., spontaneous eye-opening), an observation that was corroborated by similarly fast changes in EEG signals. Collectively, these results provide the basis for further investigation into IV MPH as a safe therapy that may rapidly upregulate brain networks involved in brain injury recovery.

The proof-of-principle findings in this Phase 1 study address a critical gap in clinical care by demonstrating that it may be feasible and safe to pharmacologically promote early recovery of arousal, a critical component of consciousness, in ICU patients with acute severe TBI. Currently, there are no therapies approved by the United States FDA for this purpose, though encouraging results from pharmacologic^6^ and device-based^34^ clinical trials highlight the potential to upregulate neural networks and restore consciousness in the acute stage of severe TBI recovery. Our observations using the dopamine reuptake inhibitor IV MPH are consistent with a recent acute study of subcutaneous apomorphine and enteral methylphenidate,^6^ and a Phase 3 study of amantadine hydrochloride in the subacute setting (4 to 16 weeks post-injury),^7^ as each of these studies indicates a key role for dopamine neurotransmission in reengaging brain networks essential for consciousness. We extend these prior findings in two ways: first, by demonstrating the benefit of IV administration of dopaminergic therapy, as IV MPH was found to have a rapid T_max_ (7 minutes at the 0.5 mg/kg dose); and second, by using complementary EEG and rs-fMRI pharmacodynamic analyses to demonstrate rapid target engagement, as pharmacodynamic responses in some participants were observed via both modalities within minutes of IV MPH bolus administration.

In considering dose selection for future Phase 2 trials, all three doses of IV MPH tested here – 0.5, 1.0, and 2.0 mg/kg – met our prespecified criteria for future testing: all were associated with fewer than 2 drug-related SAEs, and all were associated with a ≥10% rate of pharmacodynamic responses on EEG or rs-fMRI.^29^ Notably, the former safety criterion was based upon a target enrollment of 22 participants, highlighting the importance of acquiring additional safety data in Phase 2. The high rate of pharmacodynamic responses observed on EEG, which generally exceeded the rate of behavioral improvements on the CRS-R, suggests that IV MPH may initiate subclinical changes in brain function that evade or precede detection on bedside behavioral examination. Though only two participants underwent rs-fMRI pharmacodynamic analysis, the rs-fMRI findings provide complementary evidence, in a single participant, that IV MPH can reactivate cortical networks involved in conscious awareness (i.e., the DMN) via reintegration of dopaminergic VTA-DMN signaling. Our findings with IV MPH add to growing evidence from studies of enteral MPH^21^ that ascending mesocortical projections from dopaminergic VTA neurons play a key role in the restoration of consciousness after brain injury.

The behavioral findings in this Phase 1 study must be interpreted with caution, given its open-label design. Nevertheless, the rapid emergence of spontaneous eye-opening after IV MPH – an objective observation that is less susceptible to examiner bias than ascriptions of purposeful behavior – provides a behavioral correlate for the pharmacodynamic observations made with EEG and rs-fMRI. The high rate of arousal improvement (67% at the 0.5 mg/kg dose) indicates that IV MPH can engage the subcortical ascending arousal network even in the presence of traumatic brainstem injury (i.e., grade 3 diffuse axonal injury).^35^ Furthermore, the rapidity of eye-opening, within 15 minutes of IV MPH bolus completion, matches the rapid T_max_ observed with pharmacokinetic analysis, as well as the timeline of ADR change-points observed with EEG and VTA-DMN connectivity changepoints observed with rs-fMRI. These pharmacokinetic and pharmacodynamic data thus point toward an ability of IV MPH to rapidly engage subcortical networks responsible for arousal, in some participants resulting in broad cortical activation, as evidenced by increases in ADR on EEG, DMN connectivity on rs-fMRI, and purposeful behavior on the CRS-R.

### Limitations

A key limitation of this Phase 1 study was its small sample size, as enrollment was impacted by supply chain disruptions, the COVID-19 pandemic, implementation of updated compounding workstreams, and a high number of excluded patients. Exclusion of patients whose post-resuscitation GCS was > 8 limits the generalizability of the results, as some individuals with GCS > 8 meet criteria for DoC^36^ and some experience a decline in level of consciousness after admission. While no SAEs were observed at any dose, our sample size only allowed us to determine that the probability of SAEs was less than 34%, 34%, and 46% at the 0.5 mg/kg, 1.0 mg/kg, and 2.0 mg/kg doses, respectively. Despite this limited statistical power to detect SAEs, our findings in patients with acute severe TBI are consistent with prior studies suggesting that IV MPH is safe to use in patients with acute coma from barbiturate overdose^37–39^ and patients with other medical conditions.^19^ We had a small number of participants who underwent pharmacodynamic testing with rs-fMRI (n=2), reflecting the logistical and safety barriers associated with performing advanced MRI studies in critically ill patients with acute severe brain injuries, for whom the presence of agitation, elevated intracranial pressure, or other medical comorbidities often precludes MRI. There is thus a need for larger studies to elucidate the mechanisms by which IV MPH engages mesocortical pathways that mediate arousal and their functional connections with cortical networks like the DMN that mediate awareness.

Another limitation is that we cannot exclude a dose-stacking effect, whereby the 1.0 mg/kg and 2.0 mg/kg doses may have had less effect on pharmacodynamic biomarkers because of lingering effects of the 0.5 mg/kg dose. Even if plasma levels of MPH returned to a pre-bolus baseline prior to each dose, indicating elimination of MPH from the plasma within 24 hours, it is possible that MPH effects on target brain networks persisted beyond plasma clearance, thereby confounding the pharmacodynamic analyses at subsequent doses. Acknowledging this limitation in the study design, there was no indication that higher doses of IV MPH (1.0 and 2.0 mg/kg) had a greater impact on target brain networks than did the 0.5 mg/kg dose. It is also important to interpret the pharmacodynamic results in the context of the fluctuations in level of consciousness that are common in critically ill patients with acute severe TBI, as reflected in the changing DoC diagnosis between Day 0 and pre-dose Day 1 for 3 of 9 patients. Future studies will need to further control for these baseline fluctuations in level of consciousness.

### Conclusions and future directions

In summary, we report preliminary evidence in a Phase 1 open-label, dose-finding study that IV MPH at doses of 0.5 to 2.0 mg/kg is safe to administer to patients with acute severe TBI in the ICU. Further, we provide proof-of-principle that IV MPH rapidly engages target brain networks that modulate human consciousness, based on complementary EEG and rs-fMRI biomarkers, as well as behavioral signs of increased arousal. While all of the doses tested here appear to be safe for future clinical trials, our results indicate that 0.5 mg/kg may be the most appropriate dose to use in Phase 2 given that no AEs were observed at 0.5 mg/kg and that pharmacodynamic responses were similar across all three doses. These findings set the stage for blinded, placebo-controlled studies that test the potential of IV MPH to stimulate early recovery of consciousness, increase prognostic accuracy, and reduce premature withdrawal of life-sustaining treatment for patients with acute severe TBI in the ICU.

## Data availability

The pharmacokinetic, pharmacodynamic, and behavioral data are available upon reasonable request to the corresponding author.

## Acknowledgments

The study was funded by the NIH Director’s Office (DP2HD101400), National Institute of Neurological Disorders and Stroke (R21NS109627, RF1NS128961, R01NS130119), National Institute of Biomedical Imaging and Bioengineering (R01EB023281, R01EB033773), National Institute on Disability, Independent Living and Rehabilitation Research (90DPTB0027), James S. McDonnell Foundation, Rappaport Foundation, Chen Institute MGH Research Scholar Award, Mass General Brigham Neuroscience Transformative Scholar Award, Mass General Brigham Radiology Innovation Award, Barbara Epstein Foundation, Inc., and the MIT/MGH Brain Arousal State Control Innovation Center (BASCIC) project. We acknowledge support from the National Institute of Neurological Disorders and Stroke Clinical Trial Methodology Course (R25NS088248) and the Harvard Catalyst | The Harvard Clinical and Translational Science Center (National Center for Advancing Translational Sciences, National Institutes of Health Award UL 1TR002541). This research was also funded in part by the Swiss National Science Foundation (P500PM 210834). The content is solely the responsibility of the authors and does not necessarily represent the official views of Harvard Catalyst, Harvard University and its affiliated academic healthcare centers, or the National Institutes of Health. We thank the members of the Patient and Family Advisory Board of the Massachusetts General Hospital Laboratory for NeuroImaging of Coma and Consciousness for their feedback and insights regarding the ethical conduct of this clinical trial. We thank Sarah Pyle for artistic design assistance with Figure 1. During manuscript preparation, we used Claude (Anthropic) to assist with minor editorial refinement of language in the abstract and introduction. The authors take full responsibility for all content.

