## Supplementary Text & Figures for "Intravenous methylphenidate for acute traumatic disorders of consciousness: A phase 1 dose-finding and target engagement study"

Yelena G. Bodien^1,13^

* contributed equally and share first authorship

^1^ Center for Neurotechnology and Neurorecovery, Department of Neurology, Massachusetts General Hospital, Harvard Medical School, Boston, MA

^2^Athinoula A. Martinos Center for Biomedical Imaging, Massachusetts General Hospital, Charlestown, MA

^3^ Department of Pharmacy, Massachusetts General Hospital, Boston, MA

^4^ Department of Brain and Cognitive Sciences, Massachusetts Institute of Technology, Cambridge, MA

^5^ Department of Medicine, Massachusetts General Hospital, Boston, MA

^6^ Department of Neurology and Neurological Sciences, Stanford School of Medicine, Stanford, CA

^7^ Department of Anesthesia, Critical Care and Pain Medicine, Massachusetts General Hospital, Boston, MA

^8^ The Picower Institute for Learning and Memory, Massachusetts Institute of Technology, Cambridge, MA

^9^ Department of Neurology, Northwestern University Feinberg School of Medicine, Chicago, IL

^10^ Division of Medical Ethics and Consortium for the Advanced Study of Brain Injury (CASBI), Weill Cornell Medical College, New York, NY

^11^ The Rockefeller University

^12^ Solomon Center for Health Law and Policy, Yale Law School, New Haven, CT

^13^ Department of Physical Medicine and Rehabilitation, Spaulding Rehabilitation Hospital, Boston, MA

^14^ School of Engineering and Carney Institute for Brain Science, Brown University, Providence, RI

^15^ Veterans Affairs RR&D Center for Neurorestoration and Neurotechnology, VA Medical Center, Providence, RI

**Correspondence to:**

Dr. Brian Edlow

Center for Neurotechnology and Neurorecovery

Massachusetts General Hospital

101 Merrimac Street – Suite 310

Boston, MA 02114, USA

**Supplementary Methods**

*Study recruitment timeline*

The study was initially on hold after Institutional Review Board approval due to lack of drug availability and updating of drug manufacturing protocols to comply with new United States and Massachusetts legislature requirements. New pharmacy requirements were met by March 20, 2020, but study launch was delayed until August, 2020 due to the COVID-19 pandemic and an institution-wide hold on clinical trials that were considered not life-saving or disease-altering. There was another pause in enrollment from November 1, 2021 to May 3, 2022 due to disruption of the MPH supply chain.

*EEG data acquisition, pre-processing, and analysis*

EEG data were acquired at sampling rates of either 256 or 512 Hz (XLTEK EEG system, Natus Medical Inc.). Post-recording, data was exported to an EDF file and imported to MATLAB (MathWorks, Natick, MA). For pre-processing and analysis, we used EEGLAB (versions 2024.2 to 2025.0.0, eeglab.org) and customized MATLAB code.

Identification of a 5-min pre-bolus baseline and EEG timestamp verification for start and completion of IV MPH bolus administration were performed manually by review of EEG-synchronized video and audio recordings in the XLTEK EEG system. We then extracted the EEG data from the beginning of the baseline to the 15 minutes after completion of the IV MPH bolus. Second, we applied a 1 Hz high-pass filter and a 60 Hz band-stop filter (both butterworth 3^rd^ order, zero-phase shift). Continuous data were transformed into 3-second epochs. Upon visual inspection, a maximum of 3 channels were removed because of frequent artefacts and noise. Subsequently, data from artefact-rich epochs were manually rejected for all the remaining channels. We then re-referenced data to the average and performed the EEGLAB in-built ICA (*runica*). Upon visual inspection of the component signals, epochs with significant artefacts were manually removed and ICA recalculated. If present, we removed eye movement and muscle artefact components. Data was low-pass filtered at 30 Hz (butterworth 3^rd^ order, zero-phase shift), re-referenced using the Hjorth Laplacian transform to optimize spatial localization and to avoid contaminating activity at the reference. Recordings with 512 Hz were downsampled to 256 Hz (*decimate*). Power spectral density was calculated for each bin and channel using the multitaper method (TW: time-bandwith product = 1, L: tapers = 2) and the Chronux toolbox.^1^ The alpha-delta ratio (ADR; alpha: 8–12 Hz, delta: 1–4 Hz) was calculated on a single-channel level and then averaged across channels.

To test for an EEG response to IV MPH, we applied a changepoint analysis^2^ using the MATLAB changepoint function (*findchangepts*). The analysis was performed separately for each participant and study day with an IV MPH bolus. Identification of changepoints was based on the ADR signal (mean ADR across channels) from the beginning of the pre-bolus baseline up to 15-min post-bolus completion, MaxNumChanges=2, and statistic="mean". The EEG response to IV MPH on a study day was considered to be positive if the first change point between initiation of the IV MPH bolus and 15 minutes post-bolus completion was positive (i.e., increase in the ADR following the change point). The EEG response to IV MPH was considered to be negative if the first change point between initiation of the IV MPH bolus and 15 minutes post-bolus completion was negative or if there were no change points during this observation window. The reason for limiting the changepoint analysis to 15 minutes post-bolus completion is that a behavioral assessment with the CRS-R was performed starting at post-bolus completion minute 15, and the arousal facilitation protocol in the CRS-R could confound an EEG analysis of the effects of IV MPH. We assessed for change points during IV MPH bolus administration based on prior studies suggesting that the rapid effects of IV MPH in the human brain could be observed within minutes of administration.^3^

In the secondary analysis, we compared the ADR between the 5-minute pre-bolus baseline and the period from the start of the IV MPH bolus to 15 min after IV MPH bolus completion. This comparison was performed for each participant and IV MPH administration day. To test for a significant change in the ADR between baseline and post-IV MPH administration, we performed a circular block bootstrap^4^ with a block length of 5 and 10^6^ iterations. The ADR of the baseline and IV MPH post-bolus periods were resampled separately in consecutive blocks to preserve short-range temporal dependence between neighboring ADR values (one ADR data point per 3 s EEG epoch). The 95% confidence interval of the bootstrapped median difference between the two conditions was then used to determine significance, with intervals excluding zero indicating a significant change of the ADR (increase or decrease) in relation to the IV MPH bolus.

*Resting-state Functional MRI (rs-fMRI) data acquisition, pre-processing, and analysis*

MRI data were acquired with a 32-channel head coil on a 3-Tesla Skyra MRI scanner (Siemens Healthineers; Erlangen, Germany) located in the Massachusetts General Hospital Neurosciences ICU. The parameters of the BOLD fMRI sequence were: echo time (TE)=30ms, repetition time (TR)=1250ms, in-plane resolution=2.0x2.0mm, slice thickness=2 mm, interslice gap=0mm, matrix=106x105, field of view=212x212mm^2^, 72 slices, simultaneous multislice (SMS) factor=4. The rs-fMRI sequence was 10 mins 22 secs long (10 mins of analyzed data). High-spatial resolution 3D T1-weighted multi-echo magnetization prepared gradient echo (MEMPRAGE) anatomical images were acquired for registration purposes, as previously described.^5^

Rs-fMRI data were analyzed using CONN^6^, and preprocessing steps included realignment with correction of susceptibility distortion interactions, slice timing correction, outlier detection, direct segmentation and MNI-space normalization, and smoothing.

For rs-fMRI functional connectivity analysis, we estimated seed-based connectivity maps and ROI-to-ROI connectivity matrices to characterize patterns of functional connectivity within the DMN, and between cortical DMN nodes and VTA. Functional connectivity strength was measured using Fisher-transformed bivariate Pearson correlation coefficients from a weighted general linear model (weighted-GLM).^6,7^ Group-level analyses were performed using a GLM. For each individual voxel a separate GLM was estimated. Inferences were performed at the level of individual clusters and based on parametric statistics from Gaussian Random Field theory.^8^ Results were thresholded using a combination of a cluster-forming p<0.001 voxel-level threshold, and a FDR-corrected p<0.05 cluster-size threshold.

**Supplementary Figure 1: Schematic Illustrating the Sequence of Data Collection and Analysis Relative to Intravenous Methylphenidate (IV MPH) administration.** Post IV-MPH blood sample data collection began after completion of the bolus. Pharmacokinetic modeling of MPH concentrations in the plasma began at the start of the bolus, leveraging the availability of a pre-bolus blood sample. Note that the timeline is not to scale but is intended to show the relationship between data acquisition and analytic windows of observation. Abbreviations: CRS-R = Coma Recovery Scale-Revised; EEG = electroencephalography; rs-fMRI = resting-state functional MRI.

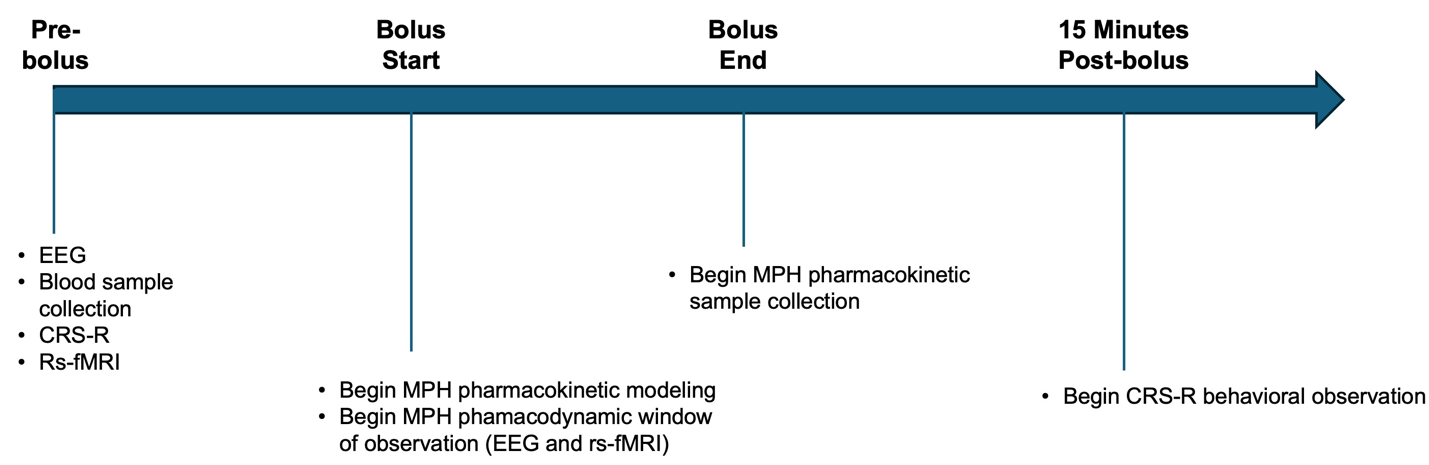

**Supplementary Figure 2. Participant Screening and Enrollment CONSORT Diagram**

Patients admitted to the MGB Neurosciences ICU between 8/24/20 – 4/1/24 were consecutively screened for enrollment. Screening was paused on 2 occasions due to factors related to intravenous methylphenidate (IV MPH) manufacturing and regulations. Abbreviations: DoC = disorders of consciousness; GCS = Glasgow Coma Scale; eMCS = emerged from the minimally conscious state; ICU = intensive care unit; PI = Principal Investigator; WLST = withdrawal of life-sustaining treatment.

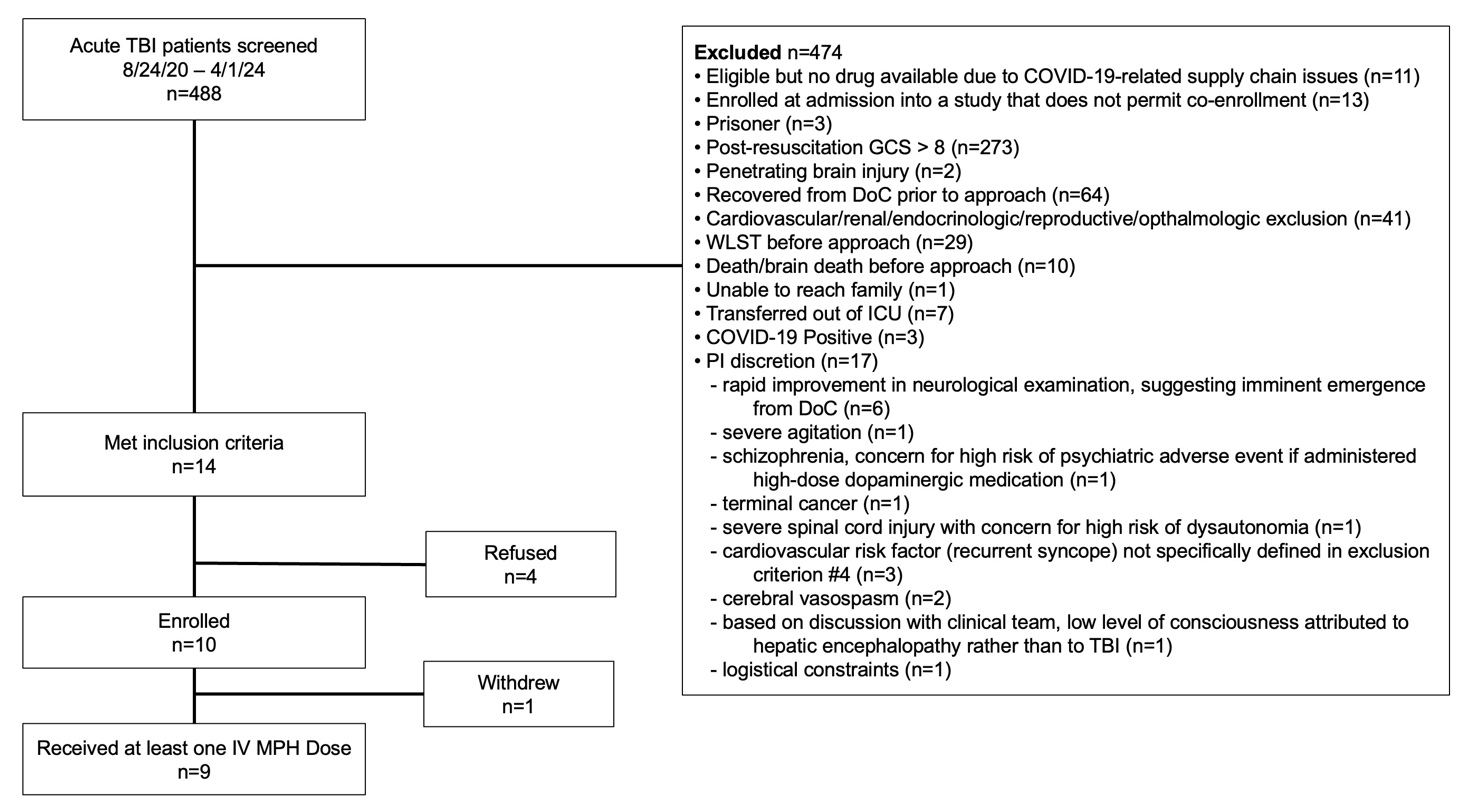

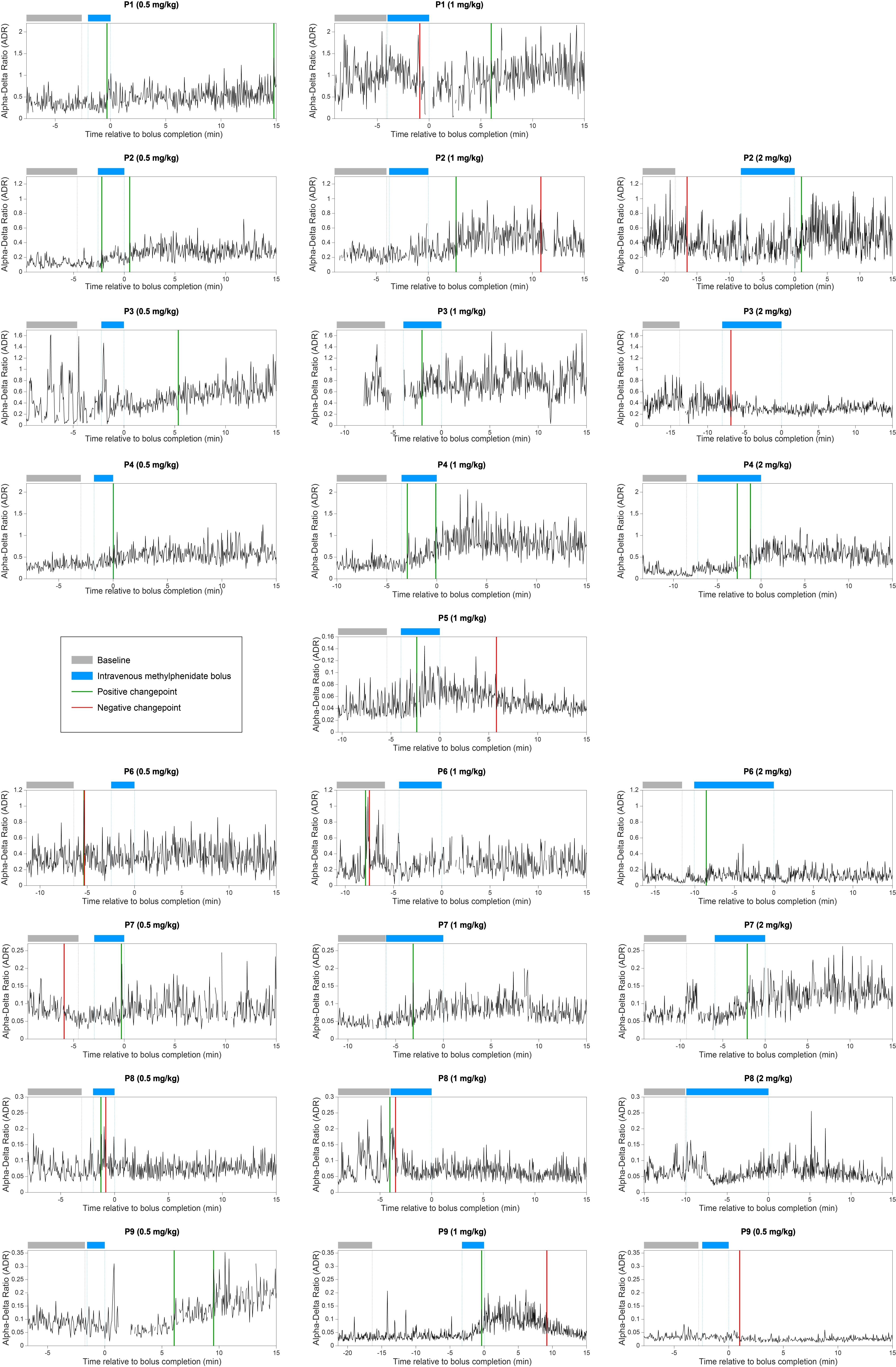

**Supplementary Figure 3: Individual Pharmacodynamic Responses to Intravenous Methylphenidate (IV MPH) Measured by Electroencephalography (EEG).** The alpha-delta ratio (ADR) measured by EEG for each patient is plotted against time. The bolus time window is indicated by a blue bar, and the pre-bolus window of ADR measurement is indicated by a grey bar. The time window for assessing an EEG pharmacodynamic response to IV MPH ends at 15 minutes, which is when a behavioral examination with the Coma Recovery Scale-Revised was performed.
