## Supplementary Tables for "Intravenous methylphenidate for acute traumatic disorders of consciousness: A phase 1 dose-finding and target engagement study"

| **ID** | **Day 0**  **Baseline** | | | **Day 1**  **Pre-IV MPH** | | |
| --- | --- | --- | --- | --- | --- | --- |
|  | **LoC** | **CRS-R Total Score** | **CRS-R**  **Subscale Scores** | **LoC** | **CRS-R Total Score** | **CRS-R**  **Subscale Scores** |
| P1 | MCS+ | 14 | A3V3M5O1C0Ar2 | MCS+ | **12** | **A4**V3**M3**O1C0**Ar1** |
| P2 | MCS- | 5 | A0V0M5O0C0Ar0 | MCS- | **6** | A0V0M5O0C0**Ar1** |
| P3 | VS/UWS | 5 | A0V0M2O1C0Ar2 | VS/UWS | **4** | A0V0M2O1C0**Ar1** |
| P4 | VS/UWS | 4 | A0V0M2O1C0Ar1 | **Coma** | **3** | A0V0M2O1C0**Ar0** |
| P5 | N/A* | N/A | N/A | MCS- | 4 | A0V0M3O0C0Ar1 |
| P6 | VS/UWS | 5 | A1V0M2O1C0Ar1 | VS/UWS | **4** | **A0**V0M2O1C0Ar1 |
| P7 | Coma | 1 | A0V0M0O1C0Ar0 | **VS/UWS** | **4** | A0V0**M2**O1C0**Ar1** |
| P8 | Coma | 2 | A0V0M1O1C0Ar0 | Coma | 2 | A0V0M1O1C0Ar0 |
| P9 | VS/UWS | 3 | A0V0M1O1C0Ar1 | **Coma**** | **1** | A0V0M1**O0**C0**Ar0** |

**Supplementary Table 1: Baseline Variance in Level of Consciousness Over 24 Hours Before First Dose of Intravenous (IV) Methylphenidate (MPH).** Level of consciousness (LoC) was assessed via behavioral evaluation with the Coma Recovery Scale-Revised (CRS-R) as coma, vegetative state (VS), minimally conscious state without language function (MCS-), or minimally conscious state with language function (MCS+). The subscales for the CRS-R are Auditory Function (A), Visual Function (V), Motor Function (M), Oromotor Function (O), Communication (C), and Arousal (Ar). Bolded scores and LoC indicate a change from the prior CRS-R assessment. If multiple CRS-R evaluations were performed on Day 0, the evaluation yielding the highest LoC is reported. * Baseline CRS-R-derived behavioral data on Day 0 are not available (N/A) because IV MPH was administered immediately after informed consent was provided, for logistical reasons.

**Day 1 of IV MPH administration delayed due to paroxysmal sympathetic hyperactivity. Thus, the first dose of IV MPH was given two days after the Day 0 CRS-R assessment.

| **ID** | **Day 1** | | | | **Day 2** | | | | **Day 3** | | | | **Day 4** | | | |
| --- | --- | --- | --- | --- | --- | --- | --- | --- | --- | --- | --- | --- | --- | --- | --- | --- |
|  | **Weight-based Dose (mg/kg)** | **Dose (mg)** | **AE** | **SAE** | **Weight-based Dose** | **Dose** | **AE** | **SAE** | **Weight-based Dose (mg/kg)** | **Dose (mg)** | **AE** | **SAE** | **Weight-based Dose (mg/kg)** | **Dose (mg)** | **AE** | **SAE** |
| P1 | 0.5 | 38.5 | 0 | 0 | 1.0 | 77.5 | 1 | 0 | --- | --- | --- | --- | --- | --- | --- | --- |
| P2 | 0.5 | 46.0 | 0 | 0 | 1.0 | 92.0 | 0 | 0 | 2.0 | 184.5 | 0 | 0 | --- | --- | --- | --- |
| P3 | 0.5 | 39.5 | 0 | 0 | 1.0 | 79.0 | 0 | 0 | 2.0 | 158.0 | 0 | 0 | --- | --- | --- | --- |
| P4 | 0.5 | 34.5 | 0 | 0 | 1.0 | 69.5 | 0 | 0 | 2.0 | 139.0 | 0 | 0 | 2.0 | 139.0 | 1 | 0 |
| P5 | 0.5 | 43.0 | 0 | 0 | 1.0 | 86.5 | 0 | 0 | --- | --- | --- | --- | --- | --- | --- | --- |
| P6 | 0.5 | 49.0 | 0 | 0 | 1.0 | 98.0 | 0 | 0 | 2.0 | 196.0 | 0 | 0 | --- | --- | --- | --- |
| P7 | 0.5 | 61.3 | 0 | 0 | 1.0 | 123.0 | 0 | 0 | 2.0 | 246.0 | 0 | 0 | --- | --- | --- | --- |
| P8 | 0.5 | 40.0 | 0 | 0 | 1.0 | 80.0 | 0 | 0 | 2.0 | 160.0 | 0 | 0 | 2.0 | 160.0 | 0 | 0 |
| P9 | 0.5 | 27.5 | 0 | 0 | 1.0 | 55.0 | 2 | 0 | 0.5 | 27.5 | 0 | 0 | --- | --- | --- | --- |

**Supplementary Table 2: Overview of IV MPH Dosing and Adverse Events at Each Dose.** Abbreviations: AE = Adverse Event; SAE = Serious Adverse Event; --- = not applicable (i.e., dose not administered).

| **ID** | **Total Doses** | **Dose on Day 1 (mg/kg)** | **Dose on Day 2 (mg/kg)** | **Dose on Day 3 (mg/kg)** | **Dose on Day 4 (mg/kg)** | **Reason(s) for Doses Missed or for Cessation of Dose Escalation** |
| --- | --- | --- | --- | --- | --- | --- |
| P1 | 2 | 0.5 | 1.0 | --- | --- | Patient emerged from MCS to PTCS after 1.0 mg/kg dose on day 2 and remained in PTCS on Days 3 and 4. If he had not emerged from MCS, he would have received 0.5 mg/kg on days 3 and 4 because of AEs on Day 2. |
| P2 | 3 | 0.5 | 1.0 | 2.0 | --- | After 2.0 mg/kg dose on Day 3, patient developed agitation and nurse did not believe that patient would be able to tolerate an MRI scan safely on Day 4. Given that the patient had already received all three doses, and that an MRI scan was deemed to be contraindicated by the clinical team, the Principal Investigator decided to stop the patient’s participation in the trial. |
| P3 | 3 | 0.5 | 1.0 | 2.0 | --- | The patient had metal in his body that precluded an MRI scan. Given that safety data had already been acquired at all three doses and that no additional pharmacodynamic data (e.g. rs-fMRI) could be acquired on Day 4, the Principal Investigator decided not to administer another dose of IV MPH on Day 4. |
| P4 | 4 | 0.5 | 1.0 | 2.0 | 2.0 | Not applicable |
| P5 | 2 | 0.5 | 1.0 | --- | --- | Patient’s family decided to transition goals of care to comfort-focused care on Day 3. The family requested that the Day 3 and Day 4 doses not be administered. |
| P6 | 3 | 0.5 | 1.0 | 2.0 | --- | The patient did not tolerate lying supine for an MRI scan due to a large body habitus, such that his abdomen compressed his thoracic cavity and compromised ventilation in the supine position. After multiple attempts by the ICU nurse and respiratory therapist to identify optimal ventilatory settings, he was still not able to tolerate lying supine.  Thus, the clinical team informed the research team that it would not be safe to pursue the Day 4 MRI. |
| P7 | 3 | 0.5 | 1.0 | 2.0 | --- | No dose on day 4 because not clinically stable to travel to MRI. |
| P8 | 4 | 0.5 | 1.0 | 2.0 | 2.0 | Not applicable. |
| P9 | 3 | 0.5 | 1.0 | 0.5 | --- | The patient experienced two AEs at 1.0 mg/kg dose – paroxysmal sympathetic hyperactivity and emesis. |

**Supplementary Table 3: IV Methylphenidate Doses Received.** Abbreviations: AE = Adverse Event; --- = dose not administered.

| **ID** | **Event Type** | **Post-TBI Day #** | **MPH to Event Onset (hours)** | **Event Duration** | **Expectedness** | **Relatedness** | **Therapies Administered** | **Implications for Subsequent**  **MPH Dosing and Trial Participation** |
| --- | --- | --- | --- | --- | --- | --- | --- | --- |
| **0.5 mg/kg** | | | | | | | | |
| P4 | Increased ALT/AST | 5 | ~20 | Surpassed AE threshold after 2^nd^ 2.0 mg/kg dose (post-TBI Day 8) | Expected | Related | None | None |
| P4 | diaphoresis | 5 | immediate | minutes | Expected | Related | None | None |
| **1.0 mg/kg** | | | | | | | | |
| P1 | restlessness | 7 | 11 | 5 | Expected | Related | --- | --- |
| P4 | Increased ALT/AST | 6 | ~20 | Surpassed AE threshold after 2^nd^ 2.0 mg/kg dose (post-TBI Day 8) | Expected | Related | None | None |
| P4 | diaphoresis | 6 | immediate | minutes | Expected | Related | None | None |
| **2.0 mg/kg** | | | | | | | | |
| P2 | agitation | 11 | 1.5 | 8 | Expected | Related | Fentanyl 50 mcg IV x3  Morphine 2 mg IV  Oxycodone 10 mg PEG | Patient stopped participation after the 2.0 mg/kg dose, but not because of this AE. Rather, per protocol, the patient could have proceeded in the trial to receive 1.0 mg/kg on day 4 (the previously tolerated dose), but the clinical team stated that an MRI on day 4 was contraindicated due to safety concerns and therefore the patient did not receive subsequent doses of IV MPH. |
| P4 | Increased ALT/AST | 7 | ~20 | Surpassed AE threshold after 2^nd^ 2.0 mg/kg dose (post-TBI Day 8) | Expected | Related | None | None |
| P4 | diaphoresis | 7 | immediate | minutes | Expected | Related | None | None |
| P4 | diaphoresis | 8 | immediate | minutes | Expected | Related | None | None |

**Supplementary Table 4: Events of Clinical Interest.** Abbreviations: AE = adverse event; ALT = alanine transaminase; AST = aspartate aminotransferase; IV = intravenous; PEG = percutaneous endoscopic gastrostomy; --- = not applicable.

| **ID** | **Day 1** | | **Day 2** | | **Day 3** | | **Day 4** | |
| --- | --- | --- | --- | --- | --- | --- | --- | --- |
|  | **Pre-MPH** | **Post-MPH** | **Pre-MPH** | **Post-MPH** | **Pre-MPH** | **Post-MPH** | **Pre-MPH** | **Post-MPH** |
| P1 | Not needed | Not needed | Not needed | Not needed | --- | --- | --- | --- |
| P2 | Not needed | 5 mg IV metoprolol x2 | Esmolol gtt at 25 mcg/kg/min | Esmolol gtt at 25-50 mcg/kg/min | Esmolol gtt at 50 mcg/kg/min | Esmolol gtt at 50-100 mcg/kg/min and clevidipine gtt at 2 mg/hr | --- | --- |
| P3 | Nicardipine gtt at 7.5 mg/hr* | Nicardipine gtt at 7.5 mg/hr* | Nicardipine gtt at 2.5 mg/hr* | Nicardipine gtt at 2.5 mg/hr* | Not needed | Not needed | --- | --- |
| P4 | Nicardipine gtt at 10 mg/hr and labetalol 20 mg IV x1 | Nicardipine gtt at 10 mg/hr | Not needed | Not needed | Not needed | Not needed | Not needed | Not needed |
| P5 | Esmolol gtt at 25 mcg/kg/min and clevidipine gtt at 1 mg/hr* | Esmolol gtt at 25-300 mcg/kg/min | Not needed | Not needed | --- | --- | --- | --- |
| P6 | Clevidipine gtt at 6mg/hr* | Clevidipine gtt at 6mg/hr* and esmolol gtt at 25 mcg/kg/min** | Not needed | Not needed | Not needed | Not needed | --- | --- |
| P7 | Not needed | Esmolol gtt at 25-75 mcg/kg/min | Not needed | Esmolol gtt at 25-50 mcg/kg/min | Not needed | Esmolol gtt at 25-125 mcg/kg/min | --- | --- |
| P8 | Not needed | Esmolol gtt at 25 mcg/kg/min and clevidipine gtt at 2-14 mg/hr* | Not needed | Esmolol gtt at 25-75 mcg/kg/min and clevidipine gtt at 2-6 mg/hr* | Esmolol gtt at 12.5 mcg/kg/min and clevidipine gtt at 1 mg/hr* | Esmolol gtt at 25-75 mcg/kg/min and clevidipine gtt at 2-8 mg/hr* | Clevidipine gtt at 1 mg/hr | Esmolol gtt at 25-75 mcg/kg/min and clevidipine gtt at 2-4 mg/hr* |
| P9 | Not needed | Not needed | Not needed | Esmolol gtt at 25-200 mcg/kg/min | Not needed | Not needed | --- | --- |

**Supplementary Table 5: Pre-MPH and Post-MPH Intravenous Medications Administered to Reach Heart Rate and Blood Pressure Targets.** Per the study protocol, HR had to be <120 bpm and SBP < 180 mmHg prior to administering the IV MPH. After each dose of IV MPH, the HR target was < 120 bpm and SBP target was < 180 mmHg. Any medications used to lower HR or BP within 1 hour prior to or after the IV MPH bolus are reported here. Abbreviations: --- = no study drug administered.

* clinical goal was SBP < 160 mmHg, which is less than the study-defined target of SBP < 180 mmHg.

** clinicians decided to start esmolol gtt at HR 110bpm, which is less than the study-defined target of HR < 120 bpm

| Subject ID | Plasma Samples  (N) | C_max_ (ng/mL) | T_max_ (hr) | AUC_0-T_ (ng*h/mL) | AUC_0-inf_ (ng*h/mL) | t_½_ (hour) | CL (L/h) | Vd (L) |
| --- | --- | --- | --- | --- | --- | --- | --- | --- |
| **0.5 mg/kg** | | | | | | | | |
| P1 | 11 | 224 | 0.08 | 436.08 | 437.36 | 3.20 | 88.03 | 406.02 |
| P3 | 11 | 282 | 0.13 | 456.18 | 487.88 | 8.23 | 80.96 | 960.91 |
| P4 | 11 | 330 | 0.12 | 1633.66 | 1778.55 | 6.11 | 19.40 | 170.85 |
| P5 | 11 | 524 | 0.12 | 1435.63 | 1524.91 | 7.24 | 28.20 | 294.46 |
| P6 | 11 | 310 | 0.13 | 327.15 | 331.73 | 5.08 | 147.71 | 1082.55 |
| P7 | 11 | 418 | 0.13 | 2032.35 | 2390.92 | 9.34 | 25.72 | 346.47 |
| P8 | 9 | 269 | 0.12 | 353.27 | 354.45 | 3.24 | 112.85 | 527.60 |
| P9 | 10 | 240.2 | 0.12 | 375.80 | 377.33 | 2.44 | 72.88 | 256.27 |
| **1.0 mg/kg** | | | | | | | | |
| P1 | 10 | 316 | 0.08 | 791.56 | 814.03 | 3.28 | 95.20 | 450.48 |
| P3 | 11 | 619 | 0.07 | 1469.12 | 1524.29 | 5.31 | 51.83 | 397.18 |
| P4 | 11 | 1171 | 0.15 | 3605.41 | 3986.97 | 8.45 | 17.43 | 212.50 |
| P5 | 9 | 1148 | 0.15 | 3504.28 | 3795.12 | 4.19 | 22.79 | 137.82 |
| P6 | 11 | 569 | 0.15 | 662.85 | 674.26 | 5.04 | 145.34 | 1056.70 |
| P7 | 11 | 862 | 0.18 | 4005.69 | 4217.84 | 5.85 | 29.16 | 245.98 |
| P8 | 9 | 460 | 0.15 | 739.46 | 741.68 | 3.08 | 107.86 | 479.99 |
| P9 | 10 | 743 | 0.15 | 744.66 | 746.55 | 2.46 | 73.67 | 261.80 |
| **2.0 mg/kg** | | | | | | | | |
| P3 | 10 | 1268 | 0.23 | 3082.21 | 3325.06 | 4.71 | 47.52 | 322.89 |
| P4 | 10 | 1981 | 0.20 | 6693.38 | 7681.92 | 8.21 | 18.09 | 214.22 |
| P6 | 11 | 973 | 0.25 | 1458.53 | 1486.53 | 5.36 | 131.85 | 1019.76 |
| P7 | 9 | 1640 | 0.30 | 6401.89 | 7531.06 | 4.40 | 32.66 | 207.21 |
| P8 | 9 | 998 | 0.22 | 1606.70 | 1616.94 | 3.46 | 98.95 | 494.33 |

**Supplementary Table 6: Patient-specific Total MPH Pharmacokinetic Parameters following IV MPH Administration**. Abbreviations: AUC0-inf, area under the concentration-time curve from time 0 to infinity; AUC0-t, area under the concentration-time curve from time 0 to last quantifiable timepoint; CL, total body clearance; Cmax, maximum plasma concentration; SD, standard deviation; t1/2, terminal elimination half-life; Tmax, time to reach maximum concentration; Vd, volume of distribution. Of note, one subject (P2) was excluded from the pharmacokinetic and statistical analysis due to inaccurate sampling.

| **ID** | **Day 1** | **Day 2** | **Day 3** | **Day 4** |
| --- | --- | --- | --- | --- |
| P1 | IV Propofol gtt | IV Propofol gtt | --- | --- |
| P2 | PO Clonidine 45 min  post-bolus | none | none | --- |
| P3 | none | none | none | --- |
| P4 | IV Dexmedetomidine gtt | IV Dexmedetomidine gtt | none | IV Dexmedetomidine gtt |
| P5 | none | none | --- | --- |
| P6 | none | none | IV Dexmedetomidine gtt | --- |
| P7 | none | none | PO Clonidine 30 min pre-bolus | --- |
| P8 | IV Dexmedetomidine gtt | PO Clobazam 1 h pre-bolus | none | none |
| P9 | IV Propofol gtt, stopped 45 min pre-bolus and restarted 50 min post-bolus | IV Dexmedetomidine gtt started 50 min post-bolus;  IV Propofol gtt started 55 min post-bolus | IV Dexmedetomidine gtt | --- |

**Supplementary Table 7: Sedative Medications Administered Within 1 Hour of Intravenous Methylphenidate Bolus.** Abbreviations: gtt = continuous drip; IV = intravenous, PO = per oral; TD = transdermal; --- = no study drug administered.

| **ID** | **Post-TBI Day #** | **Dose of IV MPH (mg)** | **~15 Minutes Pre-Dose** | | | **~15 Minutes Post-Dose** | | | **~60 Minutes Post-Dose** | | |
| --- | --- | --- | --- | --- | --- | --- | --- | --- | --- | --- | --- |
|  |  |  | **LoC** | **CRS-R Total Score** | **CRS-R Subscale Scores** | **LoC** | **CRS-R Total Score** | **CRS-R Subscale Scores** | **LoC** | **CRS-R Total Score** | **CRS-R Subscale Scores** |
| P1 | 6 | 38.5 | MCS+ | 12 | A4V3M3O1C0Ar1 | MCS+ | 12 | A4V3M3O1C0Ar1 | MCS+ | 12 | A4V3M3O1C0Ar1 |
| P2 | 9 | 46.0 | MCS- | 6 | A0V0M5O0C0Ar1 | MCS- | **9** | A0**V1**M5**O1**C0**Ar2** | MCS- | **7** | A0**V0**M5O1C0**Ar1** |
| P3 | 6 | 39.5 | VS/UWS | 4 | A0V0M2O1C0Ar1 | VS/UWS | **6** | **A1V1**M2**O0**C0**Ar2** | **MCS+** | **13** | **A4V3**M2**O1C1**Ar2 |
| P4 | 5 | 34.5 | Coma | 3 | A0V0M2O1C0Ar0 | **VS/UWS** | **6** | **A1**V0M2O1C0**Ar2** | VS/UWS | **5** | **A0**V0M2O1C0Ar2 |
| P5 | 10 | 43.0 | MCS- | 4 | A0V0M3O0C0Ar1 | MCS- | **8** | A0**V1M5O1**C0Ar1 | MCS- | **9** | **A1**V1M5O1C0Ar1 |
| P6 | 10 | 49.0 | VS/UWS | 4 | A0V0M2O1C0Ar1 | VS/UWS | **5** | A0V0M2O1C0**Ar2** | VS/UWS | **4** | A0V0M2O1C0Ar1 |
| P7 | 33 | 61.3 | VS/UWS | 4 | A0V0M2O1C0Ar1 | VS/UWS | 4 | A0V0M2O1C0Ar1 | VS/UWS | 4 | A0V0M2O1C0Ar1 |
| P8 | 13 | 40.0 | Coma | 2 | A0V0M1O1C0Ar0 | VS/UWS | **4** | A0V0M1O1C0**Ar2** | VS/UWS | **3** | A0V0M1O1C0**Ar1** |
| P9 | 22 | 27.5 | Coma | 1 | A0V0M1O0C0Ar0 | **VS/UWS** | **4** | A0V0M1**O1**C0**Ar2** | VS/UWS | **3** | A0V0M1O1C0**Ar1** |
| P9* | 24 | 27.5 | Coma | 1 | A0V0M1O0C0Ar0 | **VS/UWS** | **4** | A0V0M1**O1**C0**Ar2** | VS/UWS | 4 | A0V0M1O1C0Ar2 |

**Supplementary Table 8: Behavioral Responses to 0.5 mg/kg IV Methylphenidate**. All times for the Coma Recovery Scale-Revised (CRS-R) behavioral assessments are defined time in relation to the time of bolus completion. Level of consciousness (LoC) was assessed via behavioral evaluation with the CRS-R as coma, vegetative state (VS), minimally conscious state without language function (MCS-), or minimally conscious state with language function (MCS+). The subscales for the CRS-R are Auditory Function (A), Visual Function (V), Motor Function (M), Oromotor Function (O), Communication (C), and Arousal (Ar). Bolded scores and LoC indicate a change from the prior CRS-R assessment. Abbreviations: IV MPH = intravenous methylphenidate; TBI = traumatic brain injury. *P9 received a second dose of 0.5 mg/kg after experiencing an adverse event at the 1.0 mg/kg dose.

| **ID** | **Post-TBI Day #** | **Dose of IV MPH (mg)** | **~15 Minutes Pre-Dose** | | | **~15 Minutes Post-Dose** | | | **~60 Minutes Post-Dose** | | |
| --- | --- | --- | --- | --- | --- | --- | --- | --- | --- | --- | --- |
|  |  |  | **LoC** | **CRS-R Total Score** | **CRS-R Subscale Scores** | **LoC** | **CRS-R Total Score** | **CRS-R Subscale Scores** | **LoC** | **CRS-R Total Score** | **CRS-R Subscale Scores** |
| P1 | 7 | 77.5 | MCS+ | 17 | A4V3M5O3C1Ar1 | **PTCS** | **18** | A4V3M5O3**C2**Ar1 | PTCS* | **23** | A4**V5M6**O3C2**Ar3** |
| P2 | 10 | 92.0 | MCS- | 11 | A0V4M5O1C0Ar1 | MCS- | **12** | A0V4M5O1C0**Ar2** | **MCS+** | **18** | **A3V5**M5**O2C1**Ar2 |
| P3 | 7 | 79.0 | MCS+ | 17 | A4V5M5O2C0Ar1 | MCS+ | **15** | A4**V3M3**O2**C1Ar2** | MCS+ | 15 | A4**V4**M3O2C1**Ar1** |
| P4 | 6 | 69.5 | VS/UWS | 4 | A0V0M2O1C0Ar1 | **Coma** | **3** | A0V0M2O1C0**Ar0** | **VS/UWS** | **4** | A0V0M2O1C0**Ar1** |
| P5 | 11 | 86.5 | MCS- | 6 | A0V0M5O0C0Ar1 | MCS- | **8** | A0**V1**M5**O1**C0Ar1 | MCS- | **7** | A0V1M5**O0**C0Ar1 |
| P6 | 11 | 98.0 | VS/UWS | 5 | A1V0M2O1C0Ar1 | VS/UWS | 5 | **A0**V0M2O1C0**Ar2** | VS/UWS | 5 | A0V0M2O1C0Ar2 |
| P7 | 34 | 123.0 | Coma | 3 | A0V0M2O1C0Ar0 | **VS/UWS** | **4** | A0V0M2O1C0**Ar1** | **Coma** | 4 | **A1**V0M2O1C0**Ar0** |
| P8 | 14 | 80.0 | VS/UWS | 3 | A0V0M1O1C0Ar1 | VS/UWS | **4** | A0V0M1O1C0**Ar2** | VS/UWS | **3** | A0V0M1O1C0**Ar1** |
| P9 | 23 | 55 | Coma | 2 | A0V0M1O1C0Ar0 | ** | ** | ** | Coma | 2 | A0V0M1O1C0Ar0 |

**Supplementary Table 9: Behavioral Responses to 1.0 mg/kg IV Methylphenidate**. All times for the Coma Recovery Scale-Revised (CRS-R) behavioral assessments are defined time in relation to the time of bolus completion. Level of consciousness (LoC) was assessed via behavioral evaluation with the CRS-R as coma, vegetative state/unresponsive wakefulness syndrome (VS/UWS), minimally conscious state without language function (MCS-), minimally conscious state with language function (MCS+), or post-traumatic confusional state (PTCS). The subscales for the CRS-R are Auditory Function (A), Visual Function (V), Motor Function (M), Oromotor Function (O), Communication (C), and Arousal (Ar). Bolded scores and LoC indicate a change from the prior CRS-R assessment. Abbreviations: IV MPH = intravenous methylphenidate; TBI = traumatic brain injury.

* 60-minute post-dose exam delayed for approximately 15 minutes because patient being extubated at 60 minutes.

** CRS-R not performed due to paroxysmal sympathetic hyperactivity, requiring active medical therapy (see Adverse Event Table)

| **ID** | **Post-TBI Day #** | **Dose of IV MPH (mg)** | **~15 Minutes Pre-Dose** | | | **~15 Minutes Post-Dose** | | | **~60 Minutes Post-Dose** | | |
| --- | --- | --- | --- | --- | --- | --- | --- | --- | --- | --- | --- |
|  |  |  | **LoC** | **CRS-R Total Score** | **CRS-R Subscale Scores** | **LoC** | **CRS-R Total Score** | **CRS-R Subscale Scores** | **LoC** | **CRS-R Total Score** | **CRS-R Subscale Scores** |
| P1 | --- | --- | --- | --- | --- | --- | --- | --- | --- | --- | --- |
| P2 | 11 | 184.5 | MCS- | 8 | A0V0M5O2C0Ar1 | **MCS+** | **12** | **A3**V0M5O2**C1**Ar1 | MCS+ | 12 | A3V0M5O2C1Ar1 |
| P3 | 8 | 158.0 | MCS+ | 16 | A4V4M3O2C1Ar2 | MCS+ | **17** | A4**V5**M3O2C1Ar2 | MCS+ | **16** | A4V5M3O2C1**Ar1** |
| P4 | 7 | 139.0 | VS/UWS | 4 | A0V0M2O1C0Ar1 | **Coma** | **3** | A0V0M2O1C0**Ar0** | **VS/UWS** | **4** | A0V0M2O1C0**Ar1** |
| P5 | --- | --- | --- | --- | --- | --- | --- | --- | --- | --- | --- |
| P6 | 12 | 196.0 | VS/UWS | 5 | A0V1M2O1C0Ar1 | **MCS-** | **6** | A0**V0M4**O1C0Ar1 | MCS- | **8** | A0**V0M5**O1C0**Ar2** |
| P7 | 35 | 246.0 | VS/UWS | 4 | A0V0M2O1C0Ar1 | VS/UWS | 4 | A0V0M2O1C0Ar1 | **coma** | **3** | A0V0M2O1C0**Ar0** |
| P8 | 15 | 160.0 | VS/UWS | 3 | A0V0M1O1C0Ar1 | VS/UWS | 3 | A0V0M1O1C0Ar1 | VS/UWS | 3 | A0V0M1O1C0Ar1 |
| P9 | --- | --- | --- | --- | --- | --- | --- | --- | --- | --- | --- |

**Supplementary Table 10: Behavioral Responses to 2.0 mg/kg IV Methylphenidate.** All times for the Coma Recovery Scale-Revised (CRS-R) behavioral assessments are defined time in relation to the time of bolus completion. Level of consciousness (LoC) was assessed via behavioral evaluation with the CRS-R as coma, vegetative state (VS), minimally conscious state without language function (MCS-), or minimally conscious state with language function (MCS+). The subscales for the CRS-R are Auditory Function (A), Visual Function (V), Motor Function (M), Oromotor Function (O), Communication (C), and Arousal (Ar). Bolded scores and LoC indicate a change from the prior CRS-R assessment. Abbreviations: IV MPH = intravenous methylphenidate; TBI = traumatic brain injury; --- = no study drug administered.

| **ID** | **0.5 mg/kg** | | **1.0 mg/kg** | | **2.0 mg/kg** | | |
| --- | --- | --- | --- | --- | --- | --- | --- |
|  | **EEG Responder** | **CRS-R**  **Dx Improve** | **EEG Responder** | **CRS-R**  **Dx Improve** | **EEG Responder** | **CRS-R**  **Dx Improve** | **rs-fMRI**  **Responder** |
| P1 | Y | N | N | Y | --- | --- | --- |
| P2 | Y | N | Y | Y | Y | Y | N/A |
| P3 | Y | Y | Y | N | N | N | N/A |
| P4 | Y | Y | Y | N | Y | N | Y |
| P5 | N/A | N | Y | N | --- | --- | --- |
| P6 | N | N | N | N | Y | Y | N/A |
| P7 | Y | N | Y | Y | Y | N | N/A |
| P8 | Y | Y | N | N | N | N | N |
| P9 | Y | Y | Y | N | --- | --- | --- |

**Supplementary Table 11: Comparison of Pharmacodynamic and Behavioral Responses to Intravenous Methylphenidate (IV MPH).** The methodology of the electroencephalography (EEG) and resting-state functional MRI (rs-fMRI) pharmacodynamic analyses are described in detail in the methods section of the manuscript and the Supplementary Materials. Behavioral evaluations were performed using the Coma Recovery Scale-Revised (CRS-R). A behavioral response here is defined by a change in CRS-R-derived diagnosis (Dx) within 60 minutes or receiving the bolus of IV MPH (e.g., a transition from vegetative state/unresponsive wakefulness syndrome to minimally conscious state). If a patient received a dose more than once, the results are reported for the first administration of the dose. Abbreviation: N = no; N/A = IV MPH was administered, but pharmacodynamic biomarker was not measured at this dose; Y = yes; --- = dose not administered.
